# Multi-method investigation of factors influencing amyloid onset and impairment in three cohorts

**DOI:** 10.1101/2021.12.02.21266523

**Authors:** Tobey J. Betthauser, Murat Bilgel, Rebecca L. Koscik, Bruno M. Jedynak, Yang An, Kristina A. Kellett, Abhay Moghekar, Erin M. Jonaitis, Charles K. Stone, Corinne D. Engelman, Sanjay Asthana, Bradley T. Christian, Dean F. Wong, Marilyn Albert, Susan M. Resnick, Sterling C. Johnson, the Alzheimer’s Disease Neuroimaging Initiative

**Author notes:** Correspondence to: Tobey J. Betthauser, Wisconsin Alzheimer’s Disease Research Center, University of Wisconsin-Madison, School of Medicine and Public Health, 600 Highland Avenue, K6/428, Clinical Sciences Center, Madison, WI, 53792. Data used in preparation of this article were obtained from the Alzheimer’s Disease Neuroimaging Initiative (ADNI) database (adni.loni.usc.edu). As such, the investigators within the ADNI contributed to the design and implementation of ADNI and/or provided data but did not participate in analysis or writing of this report. A complete listing of ADNI investigators can be found at: http://adni.loni.usc.edu/wp-content/uploads/how_to_apply/ADNI_Acknowledgement_List.pdf.

## Abstract

Alzheimer’s disease biomarkers are becoming increasingly important for characterizing the longitudinal course of disease, predicting the timing of clinical and cognitive symptoms, and for recruitment and treatment monitoring in clinical trials. In this work, we develop and evaluate three methods for modeling the longitudinal course of amyloid accumulation in three cohorts using amyloid PET imaging. We then use these novel approaches to investigate factors that influence the timing of amyloid onset and the timing from amyloid onset to impairment onset in the Alzheimer’s disease continuum.

Data were acquired from the Alzheimer’s Disease Neuroimaging Initiative (ADNI), the Baltimore Longitudinal Study of Aging (BLSA) and the Wisconsin Registry for Alzheimer’s Prevention (WRAP). Amyloid PET was used to assess global amyloid burden. Three methods were evaluated for modeling amyloid accumulation using 10-fold cross-validation and hold-out validation where applicable. Estimated amyloid onset age was compared across all three modeling methods and cohorts. Cox regression and accelerated failure time models were used to investigate whether sex, apolipoprotein E genotype and *e4* carriage were associated with amyloid onset age in all cohorts. Cox regression was used to investigate whether apolipoprotein E (*e4* carriage and *e3e3, e3e4, e4e4* genotypes), sex or age of amyloid onset were associated with the time from amyloid onset to impairment onset (global Clinical Dementia Rating ≥1) in a subset of 595 ADNI participants that were not impaired prior to amyloid onset.

Model prediction and estimated amyloid onset age were similar across all three amyloid modeling methods. Sex and *apolipoprotein E-e4* carriage were not associated with PET-measured amyloid accumulation rates. Apolipoprotein E genotype and *e4* carriage, but not sex, were associated with amyloid onset age such that *e4* carriers became amyloid positive at an earlier age compared to non-carriers, and greater *e4* dosage was associated with an earlier amyloid onset age. In the ADNI, *e4* carriage, being female and a later amyloid onset age were all associated with a shorter time from amyloid onset to impairment onset. The risk of impairment onset due to age of amyloid onset was nonlinear and accelerated for amyloid onset age >65. These findings demonstrate the feasibility of modeling longitudinal amyloid accumulation to enable individualized estimates of amyloid onset age from amyloid PET imaging. These estimates provide a more direct way to investigate the role of amyloid and other factors that influence the timing of clinical impairment in Alzheimer’s disease.

## Introduction

Alzheimer’s disease is characterized by the aggregation of beta-amyloid plaques and neurofibrillary tau tangles, followed by subsequent neurodegeneration and progressive cognitive decline.^1-4^ The disease course consists of an extended “preclinical” phase wherein Alzheimer’s disease pathology is accumulating prior to the onset of clinically observable cognitive deficits.^5^ It is estimated that the preclinical phase extends as much as 20 years prior to clinical symptoms onset, though the empirical basis for this is largely at the group level^6-9^ where studies have observed that change in amyloid levels are predictable.^7, 9-11^ Robust methods for estimating amyloid onset and thus demarcating the onset of preclinical Alzheimer’s disease at the individual level are needed and now appear to be feasible.^11-15^ Although these approaches are conceptually promising, further study is needed to optimize the methodology and elucidate the timing of events and dementia risk in Alzheimer’s disease. Here we report two new autonomous methods for estimating individualized amyloid onset age and evaluate these methods together with our previously published approach.^13^ We then employ these methods to investigate factors that may influence amyloid onset age, amyloid accumulation rates and time from amyloid onset to impairment.

*Apolipoprotein E (APOE)* genotype, specifically *APOE-e4* allele carrier status and gene dosage, is the most established risk factor for beta-amyloid accumulation and cognitive impairment in sporadic Alzheimer’s disease (see review^16^). Studies investigating dementia prevalence demonstrated a gene-dose dependent relationship wherein dementia risk increases and age of dementia onset decreases with lower *APOE-e2* dosage and higher *e4* dosage.^17-19^ Additionally, some studies have observed an interaction between *APOE* and sex such that females with one or more *APOE-e4* alleles experience earlier dementia onsetrelative to *e4* non-carriers, but a similar result was not observed for male *APOE-e4* carriers.^20^ Biomarker studies have shown amyloid positivity (A+) prevalence increases with increasing *APOE-e4* dosage, and decreases with increasing *e2* gene dosage (except *e2e4* carriers) compared to *e3* homozygotes.^8, 21^ Neuroimaging studies have also shown *e4* carriers become A+ earlier than non-carriers.^8, 12, 22, 23^ Thus, a possible explanation for *e4* carriers becoming impaired earlier in life is that they start accumulating beta-amyloid pathology earlier in life. To test this hypothesis, we need to determine if the relationship between *APOE* genotype and A+ prevalence is due to an earlier amyloid onset age, a difference in amyloid plaque accumulation rates, or possibly a combination of both. There is also a need to determine whether factors like *APOE* genotype, sex, or age of amyloid onset affect the time from amyloid onset to impairment onset.

This work builds on two prior papers where we demonstrated methods that provide individualized estimates of A+ onset age based on one or more [^11^C]Pittsburgh Compound B (PiB) PET scans. These methods were developed separately in the Baltimore Longitudinal Study on Aging (BLSA)^12^ and the Wisconsin Registry for Alzheimer’s Prevention (WRAP).^13^ The BLSA study used a nonlinear mixed-effects model with random effects to derive an estimated amyloid onset age (EAOA), and showed asymptomatic *APOE-*e4 carriers exhibited earlier EAOA compared with non-carriers. In the WRAP study, group-based trajectory modeling (GBTM) and Bayes’ theorem were used to estimate EAOA and showed EAOA could be used to calculate the duration of A+ (age at procedure minus EAOA; termed amyloid chronicity). Higher amyloid chronicity was associated with faster rates of cognitive decline, increased risk of cognitive impairment, and was more strongly associated with MK-6240 tau PET levels than age in initially unimpaired WRAP participants.

In this work, we present a simplification to the GBTM approach and introduce two new autonomous methods for modeling the longitudinal time course of PET-measured beta-amyloid accumulation. The three algorithms were evaluated in three cohorts using two amyloid PET radiotracers: the Alzheimer’s Disease Neuroimaging Initiative (ADNI) using [^18^F]Florbetapir, the BLSA using [^11^C]PiB, and the WRAP using [^11^C]PiB. Our study aims were to: 1) cross-validate three methods for modeling amyloid accumulation patterns, estimating age of A+ onset and predicting past and/or future amyloid PET levels; 2) characterize associations of *APOE* genotype and sex with EAOA and amyloid accumulation rates; and 3) characterize dementia risk and time from EAOA to dementia associated with *APOE* genotype, EAOA, and sex.

## Materials and methods

### Study Participants

Data were analyzed from ADNI, BLSA, and WRAPstudies. Detailed participant demographics are shown in Table 1. Study design and information for each cohort can be found in the following references.^24-27^ In addition to each cohort’s inclusion/exclusion criteria, participants were included in this analysis if they had at least one processed amyloid PET scan and a cognitive diagnosis available. Participants’ written consent was obtained in each of the source studies according to the *Declaration of Helsinki* and under local Institutional Review Board approvals.

**Table 1.** Demographics and Cohort Summaries.

|  | **ADNI**<br>**All**<br>**Participants** | **BLSA**<br>**All**<br>**Participants** | **WRAP**<br>**All**<br>**Participants** | **ADNI**<br>**Modeling**<br>**Set** | **BLSA**<br>**Modeling**<br>**Set** | **WRAP**<br>**Modeling**<br>**Set** |
| --- | --- | --- | --- | --- | --- | --- |
| **N** | 1215 | 207 | 272 | 739 | 142 | 179 |
| **Female, n (%)** | 570 (46.9) | 104 (50.2) | 185 (68.0) | 345 (46.7) | 71 (50.0) | 120 (67.0) |
| **Race, n (%)** |  |  |  |  |  |  |
| American Indian or Alaskan Native | 2 (0.2) | 0 | 4 (1.5) | 2 (0.3) | 0 | 1 (0.6) |
| Asian | 19 (1.6) | 4 (1.9) | 1 (0.4) | 11 (1.5) | 2 (1.4) | 1 (0.6) |
| Native Hawaiian or Other Pacific Islander | 2 (0.2) | 3 (1.4) | 0 | 1 (0.1) | 2 (1.4) | 0 |
| Black or African American | 46 (3.8) | 36 (17.4) | 10 (3.7) | 24 (3.2) | 25 (17.6) | 7 (3.9) |
| White | 1127 (92.8) | 162 (78.3) | 256 (94.1) | 689 (93.2) | 112 (78.9) | 169 (94.4) |
| Other or Multiple | 16 (1.3) | 2 (1.0) | 1 (0.4) | 11 (1.5) | 1 (0.7) | 1 (0.6) |
| Unavailable/Unknown | 3 (0.2) | 0 | 0 | 1 (0.1) | 0 () | 0 |
| **APOE Genotype, n (%)** |  |  |  |  |  |  |
| e2e2 | 2 (0.2) | 2 (1.0) | 0 | 1 (0.1) | 1 (0.7) | 0 |
| e2e3 | 99 (8.1) | 25 (12.1) | 30 (11.0) | 67 (9.1) | 15 (10.6) | 16 (8.9) |
| e2e4 | 24 (2.0) | 8 (3.9) | 10 (3.7) | 15 (2.0) | 6 (4.2) | 5 (2.8) |
| e3e3 | 589 (48.5) | 118 (57.0) | 132 (48.5) | 371 (50.2) | 84 (59.2) | 88 (49.2) |
| e3e4 | 394 (32.4) | 49 (23.7) | 88 (32.4) | 233 (31.5) | 33 (23.2) | 63 (35.2) |
| e4e4 | 107 (8.8) | 3 (1.4) | 10 (3.7) | 52 (7.0) | 2 (1.4) | 7 (3.9) |
| unknown | 0 | 2 (1.0) | 2 (0.7) | 0 | 1 (0.7) | 0 |
| **Baseline**<br>**Diagnosis, n (%)** |  |  |  |  |  |  |
| Unimpaired | 461 (37.9) | 207 (100) | 261 (96.0) | 303 (41.0) | 142 (100) | 177 (98.9) |
| MCI | 559 (46.0) | 0 | 9 (3.3) | 386 (52.2) | 0 | 2 (1.1) |
| Dementia | 195 (16.0) | 0 | 2 (0.7) | 50 (6.8) | 0 | 0 |
| **Baseline Age,**<br>**mean (SD)** | 73.7 (7.6) | 75.8 (8.2) | 62.4 (6.7) | 73.4 (7.4) | 75.9 (7.9) | 61.0 (6.2) |
| **PET Follow-up**<br>**duration years,**<br>**mean (SD)** | 2.52 (2.52) | 3.52 (3.6) | 4.11 (3.43) | 4.14 (1.93) | 5.13 (3.18) | 6.25 (2.11) |
| **Number of APET**<br>**scans, n (%)** |  |  |  |  |  |  |
| 1 | 476 (39.2) | 65 (31.4) | 93 (34.2) | - | - | - |
| 2 | 307 (25.3) | 39 (18.8) | 56 (20.6) | 307 (41.5) | 39 (18.8) | 56 (20.6) |
| 3 | 235 (19.3) | 38 (18.4) | 96 (35.3) | 235 (31.8) | 38 (18.4) | 96 (35.3) |
| 4 | 163 (13.4) | 25 (12.1) | 25 (9.2) | 163 (22.1) | 25 (12.1) | 25 (9.2) |
| 5 | 33 (2.7) | 14 (6.8) | 2 (0.7) | 33 (4.5) | 14 (6.8) | 2 (0.7) |
| 6 | 1 (0.1) | 13 (6.3) | 0 | 1 (0.1) | 13 (6.3) | 0 |
| 7 | 0 | 3 (1.4) | 0 | 0 | 3 (1.4) | 0 |
| 8 | 0 | 5 (2.4) | 0 | 0 | 5 (2.4) | 0 |
| 9 | 0 | 3 (1.4) | 0 | 0 | 3 (1.4) | 0 |
| 10 | 0 | 1 (0.5) | 0 | 0 | 1 (0.5) | 0 |
| 11 | 0 | 1 (0.5) | 0 | 0 | 1 (0.5) | 0 |

Columns in *italics* indicate the subset from each cohort with two or more amyloid PET scans used for longitudinal modelling ADNI = Alzheimer's Disease Neuroimaging Initiative, BLSA = Baltimore Longitudinal Study on Aging, WRAP = Wisconsin Registry for Alzheimer's Prevention, APOE = apolipoprotein, MCI = mild cognitive impairment, APET = amyloid positron emission tomography, SD = standard deviation

#### Cohort Descriptions

ADNI: Data used in the preparation of this article were obtained from the Alzheimer’s Disease Neuroimaging Initiative (ADNI) database (adni.loni.usc.edu). ADNI was launched in 2003 as a public-private partnership, led by Principal Investigator Michael W. Weiner, MD. The primary goal of ADNI has been to test whether serial MRI, PET, other biological markers, and clinical and neuropsychological assessment can be combined to measure the progression of MCI and early Alzheimer’s disease. For up-to-date information, see www.adni-info.org. Diagnostic cognitive status was established from the ADNI diagnosis table.

BLSA: At enrollment into the BLSA PET neuroimaging substudy,^27^ participants were free of CNS disease, severe cardiac disease, severe pulmonary disease, and metastatic cancer. Participants with a Clinical Dementia Rating (CDR)^28^ of zero and ≤3 errors on the Blessed Information-Memory-Concentration test^29^ were categorized as cognitively normal; otherwise, cognitive status was determined by consensus case conference upon review of clinical and neuropsychological data. Dementia and MCI diagnoses were based on Diagnostic and Statistical Manual of Mental Disorders^30^ (DSM-IIIR) and Petersen criteria,^31^ respectively.

WRAP: WRAP participants are unimpaired at enrollment and complete comprehensive neuropsychological assessments approximately biennially.^24^ Cognitive status (unimpaired, MCI, dementia) was established by consensus conference as previously reported for WRAP participants.^32^

Within each cohort, diagnostic status (unimpaired, mild cognitive impairment, dementia) was established for descriptive purposes using similar criteria. Clinical progression was assessed using the CDR scale when applicable.^28^

### Amyloid PET Processing, Quantification and Positivity

Cortical amyloid burden was assessed using either dynamic [^11^C]PiB (BLSA and WRAP) or [^18^F]Florbetapir (ADNI) PET imaging (see^33-37^ for PET protocol details) with separate processing and quantification methods from each cohort (see supplemental methods for details).^34, 35^ ADNI cortical Florbetapir standard uptake value ratios (SUVRs; eroded white matter reference region) processed by the Banner Alzheimer’s Institute were downloaded from the November 22, 2019 data freeze. Mean cortical PiB distribution volume ratios (DVRs; cerebellar gray matter reference region) were estimated using different reference tissue methods for BLSA^37^ and WRAP.^36^ A+ thresholds were established based on separate studies for each cohort. Florbetapir SUVR_WM_ >0.8 was used for ADNI based on receiver operating characteristic analyses (AUC 0.91, 91% negative agreement, 82% positive agreement) comparing the Banner Alzheimer’s Institute Florbetapir SUVR with a published threshold from Berkeley processed data.^38^ BLSA used mean cortical PiB DVR>1.066 derived from Gaussian mixture modeling.^12^ Global PiB DVR >1.19 was used for WRAP based on receiver operating characteristic analysis with visual assessment as the reference standard.^39^

### Amyloid Trajectory Modeling and Estimated Amyloid Onset Age (EAOA)

Three methods were used to model longitudinal amyloid PET trajectories and estimate EAOA. A brief description of each method is provided below with additional details included in the supplement. For all analyses, models were trained separately for each cohort using subsets of participants with ≥2 longitudinal amyloid PET scans (table 1, right three columns). Trained models were then used to estimate the age of A+ onset and A+ duration (A+ duration = age at procedure – EAOA) for all participants.

#### Group-based trajectory modeling (GBTM)

The GBTM EAOA method used in this study begins as previously described^13^ by applying GBTM^40, 41^ to identify the optimal number and shape of age-related DVR/SUVR group trajectory equations (up to cubic polynomial). EAOA is calculated for each fitted group function by solving for the age each function intersects the A+ threshold. For an intercept-only function (representing the subset that is not accumulating amyloid), life expectancy at time of last scan is used as previously described^13^ to estimate EAOA for that group function. Each participant’s EAOA is then calculated as the weighted sum of group function EAOA’s where residuals for each function are used to derive the weights (see supplement for details). DVR/SUVR prediction can be accomplished by modeling the DVR/SUVR vs. A+ duration curve using piecewise regression, and then solving this modeled curve for DVR/SUVR at the time difference between the reference scan and the scan of interest for prediction.

#### Ordinary differential equation – Gaussian Process (ODE-GP)

The ODE-GP algorithm is a gradient matching method^42^ that fits a non-parametric ordinary differential equation (ODE) to data. The ODE-GP algorithm has two steps. The first step consists in estimating DVR/SUVR gradients for each participant using linear regression. The second step consists in solving a non-standard GP regression problem using the data from all participants in the sample, accounting for both the noise in the measurements and in the estimated gradients. An approximate solution is provided using a second-order Taylor expansion of the Gaussian process. We choose a standard kernel: the Gaussian kernel. The radius of the kernel is estimated by maximizing the marginal likelihood of the data. The fitted model allows for the prediction of past and future DVR/SUVR for a participant using a single DVR/SUVR value at a given age as an initial condition for the ODE. The prediction is made by integrating numerically the ODE using the Euler method. Since the fitted model is autonomous (time-invariant), we summarize the fitted data with a growth curve defining the DVR/SUVR as function of the time since the A+ threshold is achieved.

#### Sampled Iterative Local Approximation (SILA)

The SILA algorithm uses discrete sampling of DVR/SUVR vs. age data to establish the first order relationship between DVR/SUVR rate and DVR/SUVR. Numerical smoothing (robust LOESS) and Euler’s method are then used to numerically integrate these data to generate a non-parametric DVR/SUVR vs. A+ duration curve with zero time corresponding to the A+ threshold. EAOA is calculated for each person by first solving this curve for time using a person’s observed DVR/SUVR, and then subtracting the model estimated A+ duration from their age at that scan. SUVR/DVR is estimated for antecedent or prospective scans by solving the DVR/SUVR vs. A+ duration curve for DVR/SUVR at the A+ duration corresponding to the time difference between reference and target scans.

### Statistical Analyses

#### Aim 1: Evaluating and Comparing Amyloid Modeling Methods

Aim 1 compared EAOA and A+ duration outcomes across the three methods and evaluated the predictive performance of each method within each of the three cohorts. The composition of the model training and testing sets varied depending on the analysis goals as described below.

#### Inter-method comparisons of EAOA and amyloid accumulation curves

EAOA and SUVR/DVR vs. A+ duration curves were compared between all three methods in each cohort. Models were trained separately in each cohort using subsets of participants with ≥2 PET scans (Table 1, right three columns). EAOA and A+ duration were then estimated for all participants using each participant’s observed DVR/SUVR and age at their last available amyloid PET scan. EAOA was compared between methods using Pearson correlations separately for A+ and A-groups. Amyloid accumulation curves (i.e., DVR/SUVR vs. estimated A+ duration) were compared by plotting observed DVR/SUVR values at the first scan as a function of estimated A+ duration referencing the last scan for participants with ≥2 scans. EAOA accuracy was evaluated in A- to A+ converters (*n*=72 ADNI; *n*=6 BLSA; *n*=22 WRAP) by calculating the proportion of times age last A- ≤ EAOA ≤ age first A+ and calculating the conversion midpoint error (years between EAOA and the midpoint age between the last A- and first A+ observations).

#### Method predictive validity

The predictive validity for each method to predict future and past DVR/SUVR and A+/-status was evaluated using holdout validation (all methods; ADNI) and 10-fold cross-validation (ODE-GP and SILA; all cohorts) based on the practicality of implementing these approaches for each method and cohort. Stratification of 10-fold cross-validation and holdout validation samples are provided in the supplemental methods. For each method and cohort, models were trained on subsets of longitudinal data, and then DVR/SUVR and age at a single reference scan (last or first) were used to predict DVR/SUVR at an earlier or later PET scan for participants left out of model training. Forward prediction was evaluated by predicting DVR/SUVR at each person’s last observation inputting each person’s first observation whereas backward prediction used the last observation to predict DVR/SUVR at the first observation. DVR/SUVR prediction residuals were used to investigate possible associations with factors that might influence model prediction (reference DVR/SUVR, age, and time from reference scan) and to generate model summary statistics. A+/-prediction performance was evaluated using balanced accuracy (adjusted for imbalanced A+ and A- frequency), sensitivity, and specificity. We also compared EAOA estimates derived from in-sample vs. out-of-sample prediction schemes graphically and using Pearson correlations. After observing similar model performance for ODE-GP and SILA methods, analyses for aims 2 and 3 below used the average of ODE-GP and SILA EAOA.

### Aim 2: *APOE* and sex associations with EAOA and amyloid accumulation rate

The impact of sex and *APOE-e4* carriage on amyloid accumulation rates were evaluated by investigating the relationship between DVR/SUVR rate and DVR/SUVR. Smoothed DVR/SUVR rate vs. DVR/SUVR curves were generated for each cohort, and separately for comparison groups (*APOE-e4* carriers vs. non-carriers; males vs. females) by applying local regression with weighted linear least squares (LOWESS) to the mean within person DVR/SUVR rate vs. DVR/SUVR data. Ninety-five percent confidence intervals were estimated using 1000 bootstrapped samples with replacement.

The second part of aim two addressed three main questions 1) what age does A+ onset occur, 2) what is the A+ risk conferred by *APOE*, and 3) does A+ onset differ between *APOE* groups? We assessed whether and by how much EAOA differs across *APOE* genotypes using survival analyses, as previously described.^43^ The event time was EAOA for A+ individuals and age at last PET observation (right-censored) for individuals that were A- but could become A+. Kaplan-Meier plots by *APOE-e4* carriage (excluding e2e4), and separately by *APOE* genotype (*e2e3, e2e4, e3e3, e3e4*, and *e4e4*), were compared in each dataset. Survival curves were compared using log-rank tests with pairwise comparisons conducted for *APOE* groups. Median survival times with 95% confidence intervals for *APOE* groups (*e4* carriers and non-carriers; *APOE* genotypes) were reported for groups that reached 50% probability of remaining A-. *APOE-e2e2* carriers were excluded from genotype analyses due to low frequency. We used Cox proportional hazards models to quantify A+ risk by *APOE-e4* carriage, and by *APOE* genotype in separate models. We used accelerated failure time models to quantify relative differences in EAOA by *APOE*-e4 carriage and genotype. Cox proportional hazards and the accelerated failure time models included sex as a covariate.

### Aim 3: Effects of *APOE*, sex and EAOA on time from A+ onset to dementia

Due to differences in recruitment strategies, WRAP and BLSA observed few clinical conversions compared to ADNI. Therefore, we used a subset of 595 ADNI participants to investigate the effects of EAOA, sex and *APOE* on time from EAOA to impairment onset. ADNI participants were included if they were not impaired before EAOA, with CDR ≥1 as the criterion for impairment.^28^ *APOE-e2e4* carriers were excluded due to the combination of risk and protective effects, and low frequency (*n*=15). We used Cox survival models to estimate hazard and survival functions of impairment from A+ onset and their relationship with *APOE-e4* carriage, sex and EAOA. The Cox model outcome was time from EAOA to impairment for individuals who became impaired, and time from EAOA to age at last CDR (right censored) for those that remained unimpaired. Predictors included education, sex, *APOE-e4* carriage, EAOA, EAOA^2^, sex*EAOA, sex*EAOA^2^, *APOE*\*EAOA, *APOE*\*EAOA^2^. EAOA^2^ was included to allow for accelerated log hazard of impairment with increasing EAOA. Non-significant interactions were removed from the model. Analyses were repeated using *APOE* genotypes (*e3e3, e3e4, e4e4*) instead of *APOE-e4* carriage. Results are reported as hazard ratios, median survival time, and the ten-year unimpaired rate with confidence intervals estimated using 1000 bootstrapped samples with replacement.

### Data availability

Data from the WRAP (https://wrap.wisc.edu) and BLSA (https://blsa.nih.gov) can be requested through an online submission process. ADNI data can be obtained via the ADNI and LONI websites.

## Results

### Study Sample Descriptive Statistics and Demographics

Table 1 shows demographics and summary statistics for the three cohorts and the subsets used for model training. ADNI and BLSA cohorts were 11.3 and 13.4 years older, respectively, than WRAP, which had a mean age of 62.4 years at baseline amyloid PET scan. BLSA and WRAP samples were primarily unimpaired at baseline PET whereas ADNI contained 62% impaired participants (MCI or dementia). Study participants primarily identified as white (ADNI: 93%, BLSA: 78%, WRAP: 94%). Demographic variables were similar for the subsets used for model training and testing (Table 1, right three columns). The mean (SD) PET follow-up for these subsets was 4.14 (1.93), 5.13 (3.18), and 6.25 (2.11) years for ADNI, BLSA and WRAP, respectively.

### Aim 1: Amyloid modeling methods comparison and evaluation

#### Inter-method comparisons

Amyloid vs. age data and A+ thresholds are shown for each cohort in Figure 1 (top row). Each cohort had a visually apparent group of amyloid non-accumulators below the A+ threshold and a group of amyloid accumulators crossing and above the threshold. Overall, the three methods produced similar amyloid accumulation patterns (middle row Figure 1), with the largest differences observed in the ADNI dataset. EAOA was highly linearly correlated between method pairs in A+ participants (r_Pearson_ = 0.88-1.00) with lower correlations observed in A- participants (r_Pearson_ = 0.76-0.95; bottom row Figure 1; supplemental tables 2 and 3). The largest inter-method EAOA differences were observed in A- participants and at high DVR/SUVR where longitudinal observations were less frequent (supplemental figure 1). In the subsets of A-/+ convertors, EAOA accuracy ranged from 16.7% (1/6, GBTM in BLSA) to 77% (17/22, SILA in WRAP), and the mean error ranged from −3.65 (GBTM in ADNI) to 1.96 years (GBTM in BLSA) across all cohorts and methods (supplemental figure 2 and supplemental table 4). GBTM, ODE-GP, and SILA had similar EAOA accuracy and errors in BLSA and WRAP cohorts, whereas EAOA accuracy was lower, and errors were higher for GBTM in ADNI compared to ODE-GP and SILA.

**Table 2:** ADNI participant demographics for aim 3 analyses.

|  | Analysis sample | Cognitively normal during follow up | Impaired after A+ onset | P values* |
| --- | --- | --- | --- | --- |
| **N subjects** | 595 | 332 (55.8%) | 263 (44.2%) |  |
| **Males** | 327 (55.0%) | 175 (52.7%) | 152 (57.8%) | 0.22 |
| **APOE e4+** | 364 (61.2%) | 175 (52.7%) | 189 (71.9%) | <.0001 |
| **White** | 566 (95.1%) | 316 (95.2%) | 250 (95.1%) | 0.94 |
| **Education** | 16.0 (2.8)<br>6-20 | 16.1 (2.8)<br>6-20 | 15.8 (2.7)<br>8-20 | 0.18 |
| **A+ onset age** | 68.5 (8.7)<br>41.1 – 94.9 | 70.8 (8.0)<br>49.0 – 94.9 | 65.7 (8.7)<br>41.1 – 88.6 | <.0001 |
| **Time (yrs) to censor/event** | 9.4 (4.5)<br>0.2 – 24.4 | 8.5 (4.5)<br>0.2 – 20.8 | 10.4 (4.4)<br>0.4 – 24.4 | <.0001 |
| **AV45 SUVR last visit** | 1.03 (0.17)<br>0.80 – 1.96 | 0.98 (0.14)<br>0.80 – 1.59 | 1.09 (0.18)<br>0.80 – 1.96 | <.0001 |
| **Age at first CDR** | 74.1 (6.9)<br>55.0-90.3 | 74.6 (6.5)<br>55.0-90.1 | 73.5 (7.3)<br>55.1-90.3 | 0.041 |
| **Age at last CDR** | 78.6 (7.7)<br>55.7-97.4 | 79.3 (7.5)<br>59.0-97.4 | 77.7 (7.9)<br>55.7-96.0 | 0.014 |
| **Cog Follow up (Yrs)** | 4.5 (3.4)<br>0-15.1 | 4.7 (3.3)<br>0-15.1 | 4.3 (3.5)<br>0-14.8 | 0.15 |
| **AV45 last age** | 77.4 (7.6)<br>55.6 – 96.5 | 78.2 (7.3)<br>59.0 – 96.5 | 76.5 (7.9)<br>55.6 – 96.0 | 0.0061 |

Continuous variables are represented as mean (SD) with the range shown in the second line. Participants were included in aim three analyses if they were not impaired (i.e. CDR<1) before the estimated age of A+ onset. Fifteen APOE e2e4 carriers were excluded from the analyses due to small sample size (n=15) and the mixture of risk and protective alleles. Group comparisons (impaired during follow-up vs. not impaired during follow-up) for continuous and categorical variables used t-tests and chi-square tests, respectively.

**Figure 1:**
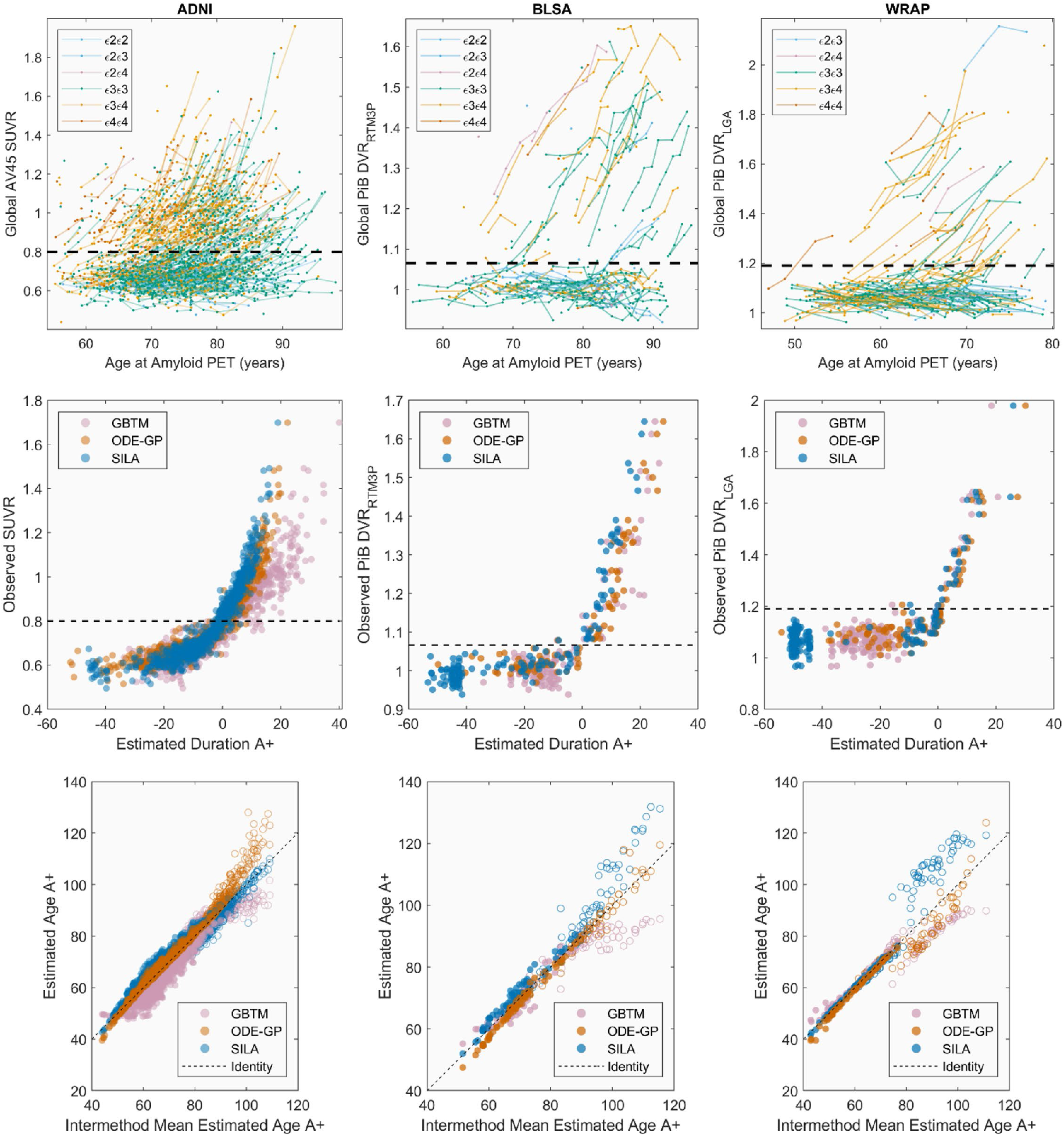
Observed and modeled amyloid trajectories and estimated amyloid onset age. Observed (top row) and amyloid accumulation patterns (middle row), and EAOA (bottom row) for each method and cohort (each column represents a cohort). APOE genotype is color coded for plots in the top row. The plots in the middle row represent backwards prediction of the duration A+ (estimated duration A+ at first scan using last scan for within-person reference). Open circles in the bottom row indicate participants who were A- at their last observation and demonstrate the instability of estimating the onset age of A+ in cases with low amyloid levels. This suggests that interpretation of EAOA in A- cases is likely limited.

#### Forward and Backward Predictive Validity

The time for prediction evaluation ranged from 0.9 to 13.1 years. Using holdout cross-validation in ADNI, all three methods had high balanced accuracy (94-98%) for predicting future or antecedent A+/-status (supplemental table 5). Sum of squared residuals were ODE-GP < SILA < GBTM for backward SUVR prediction, and ODE-GP < GBTM < SILA for forward SUVR prediction with all methods except GBTM reporting higher residual sum of squares for forward compared to backward SUVR prediction. Using 10-fold cross-validation in all cohorts, ODE-GP and SILA had similar SUVR/DVR predictive performance with balanced accuracy ranging from 94-99% for backward A+/-prediction and 82-96% for forward A+/-prediction. Prediction error was lower for backward prediction compared to forward prediction for ODE-GP and SILA in all three cohorts. Model residuals were weakly associated with age at reference scan (|Spearman rho| ≤0.26) and time to/from reference scan (|Spearman rho| ≤0.28) for all three methods and cohorts (supplemental tables 6-9, supplemental figures 3-6). ODE-GP and SILA had weak to moderate associations between prediction residuals and reference SUVR/DVR (|r_Spearman_| ≤0.41), whereas GBTM had a moderate association in the ADNI dataset (r_Spearman_ =0.45 and 0.61 for backwards and forwards prediction, respectively). EAOA was highly linearly correlated (r_Pearson_ >0.99) for all methods and cohorts when using out-of-sample (10-fold or holdout cross-validation) vs. in-sample training/prediction paradigms (supplemental Figure 7).

### Aim 2: EAOA, sex and *APOE* genotypes

#### *APOE*, sex and rate of amyloid accumulation

No differences were observed in SUVR/DVR rates vs. SUVR/DVR between males and females or between *APOE-ε4* carriers and non-carriers (Figure 2, Supplemental Figures 8-9).

**FIG 2:**
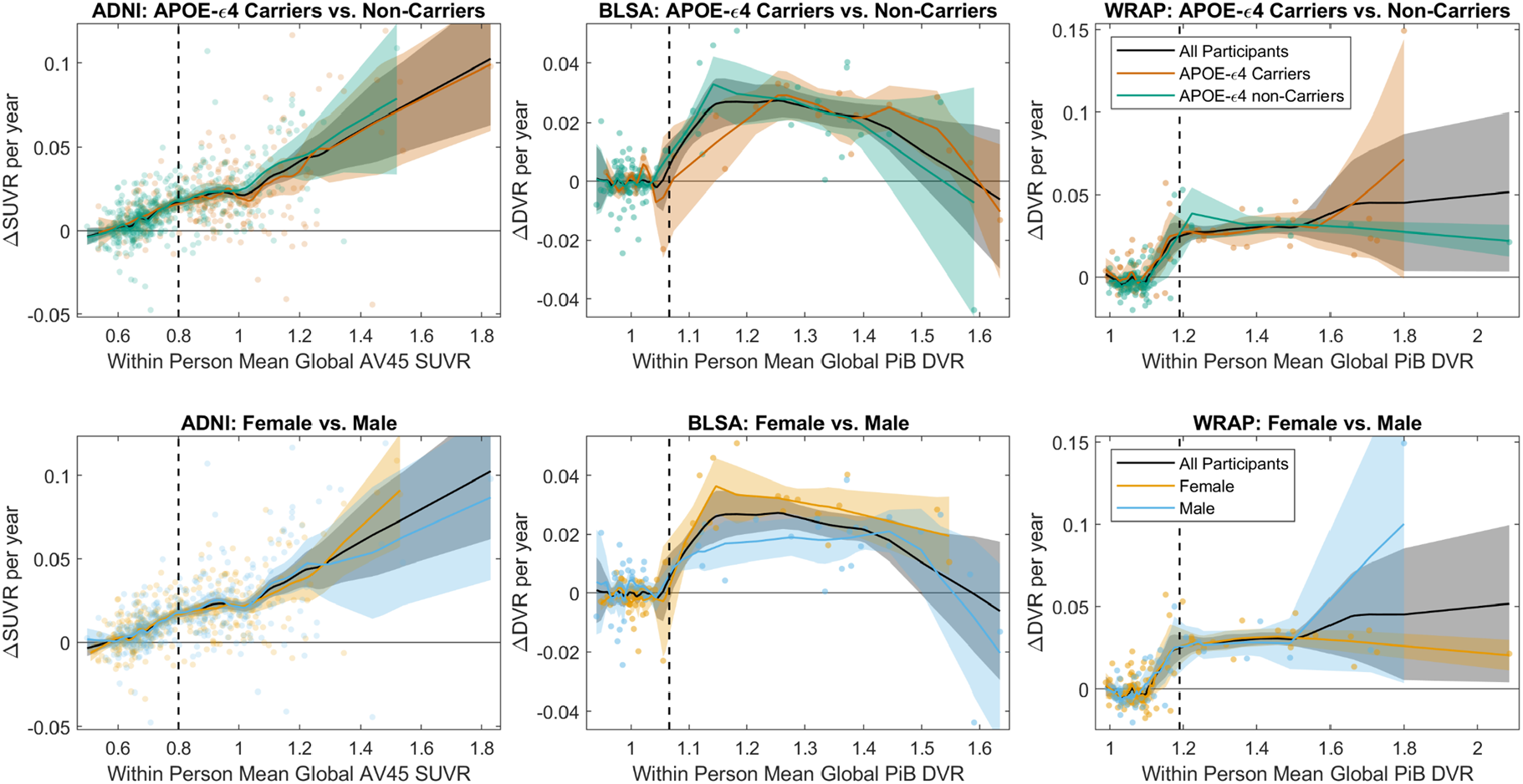
Amyloid accumulation rates by *APOE* Carriage and Sex. Comparisons of amyloid accumulation rates as a function of amyloid level between *APOE-e4* carriers and non-carriers (top row) and between males and females (bottom row) for each of the three cohorts (columns). Scatter plots show the within-person amyloid rate as a function of their mean DVR or SUVR value. Lowess lines show the average relationship between the rate of amyloid change and the amyloid level for the entire sample, and for subsets of *APOE-e4* carriers and non-carriers and for females and males. Shaded regions represent the 95% confidence intervals derived from 1000 bootstrapped samples. The relationship between the amyloid accumulation rate and the level of amyloid burden was similar between *APOE-e4* carriers vs. non-carriers, and between males vs. females across all cohorts. This suggests the rate of amyloid accumulation does not differ by *APOE-e4* carriage or sex.

#### Effect of *APOE* and sex on EAOA and A+ risk

##### What age does A+ onset occur?

EAOA by *APOE* genotype is shown for each cohort in Figure 3 for observed A+ participants. Kaplan-Meier curves by *APOE-*e4 status and genotype are presented in Figure 4. The difference in Kaplan-Meier curves between *APOE-e4-* and *e4+* (excluding *e2e4*) groups was statistically significant in each dataset (ADNI *P<*10^−15^, BLSA *P=*0.003, WRAP *P=*10^−11^). In ADNI, the median A- survival age was 84.2 (95% confidence limits: 82.7, 88.1) for the *e4-* group and 69.1 (68.2, 70.5) for the *e4*+ group. Median survival times for *e4-* could not be estimated in BLSA and WRAP because this group did not attain 50% A+ probability. Median survival time for the *e4*+ group was 83.0 (lower 95% confidence limit =72.1) in BLSA and 70.7 (67.0, 73.0) in WRAP. In ADNI, all pairwise survival curve comparisons among *e2e3, e2e4, e3e3, e3e4*, and *e4e4* groups were statistically significant (all *P<*0.0008) except for differences between *e2e4* and *e3e4* (*P*=0.33). In WRAP, *e2e3* vs. *e3e4, e2e3* vs. *e4e4, e3e3* vs. *e3e4*, and *e3e3* vs. *e4e4* were significant (all *P<*0.0002). No significant pairwise comparisons were found in BLSA, but the *e3e3* vs. *e3e4* and *e3e3* vs. *e4e4* comparisons were near significance (*P*=0.059). In ADNI, the median A- survival time was 83.1 (81.3, 85.4) for the *e3e3* group, 74.2 (lower 95% confidence limit =67.4) for *e2e4*, 70.5 (69.4, 71.8) for *e3e4*, and 64.0 (62.5, 66.4) for *e4e4*. In BLSA, the median survival time was 83.0 (lower 95% confidence limit = 72.1) for *e3e4* and 63.5 (lower 95% confidence limit =57.2) for *e4e4*, and in WRAP, 70.7 (67.7, 73.0) and 61.7 (lower 95% confidence limit =49.1), respectively.

**FIG 3.**
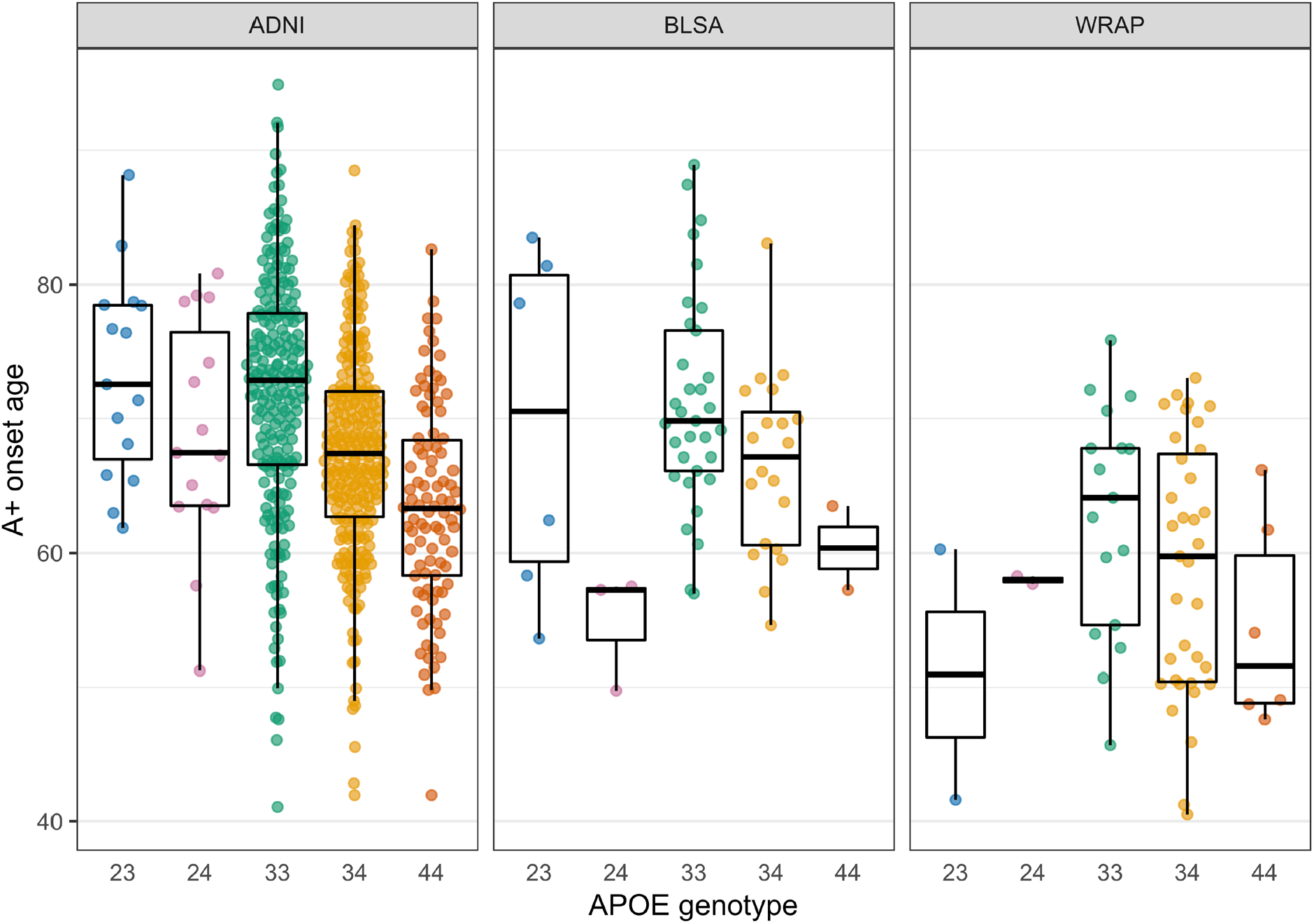
Estimated amyloid onset age by *APOE* genotype. Estimates are shown only for individuals whose observed A+ onset preceded their last PET visit. Data shown are the average of the estimates computed by the SILA and ODE-GP methods. To assess whether A+ onset age differs by *APOE* genotype, we conducted pairwise comparisons using accelerated failure time models, which allow inclusion of A- participants in the analyses via right-censoring. The e3e4 and e4e4 groups had an earlier A+ onset compared to e3e3 in all data sets. The e3e4 A+ onset age was 13%, 12%, and 20% earlier in the ADNI, BLSA, and WRAP, respectively, and the e4e4 A+ onset age was 21%, 24, and 30% earlier. The e2e4 group had an earlier amyloid onset than the e3e3 in the ADNI (10% earlier) and BLSA (19% earlier). e2e3 had a later amyloid onset compared to e3e3 in the ADNI only (13% later). The remaining comparisons with the e3e3 group were not statistically significant. in all cohorts, and additionally for the e2e4 group in ADNI. e2e3 carriers had a lower hazard in ADNI.

**FIG 4.**
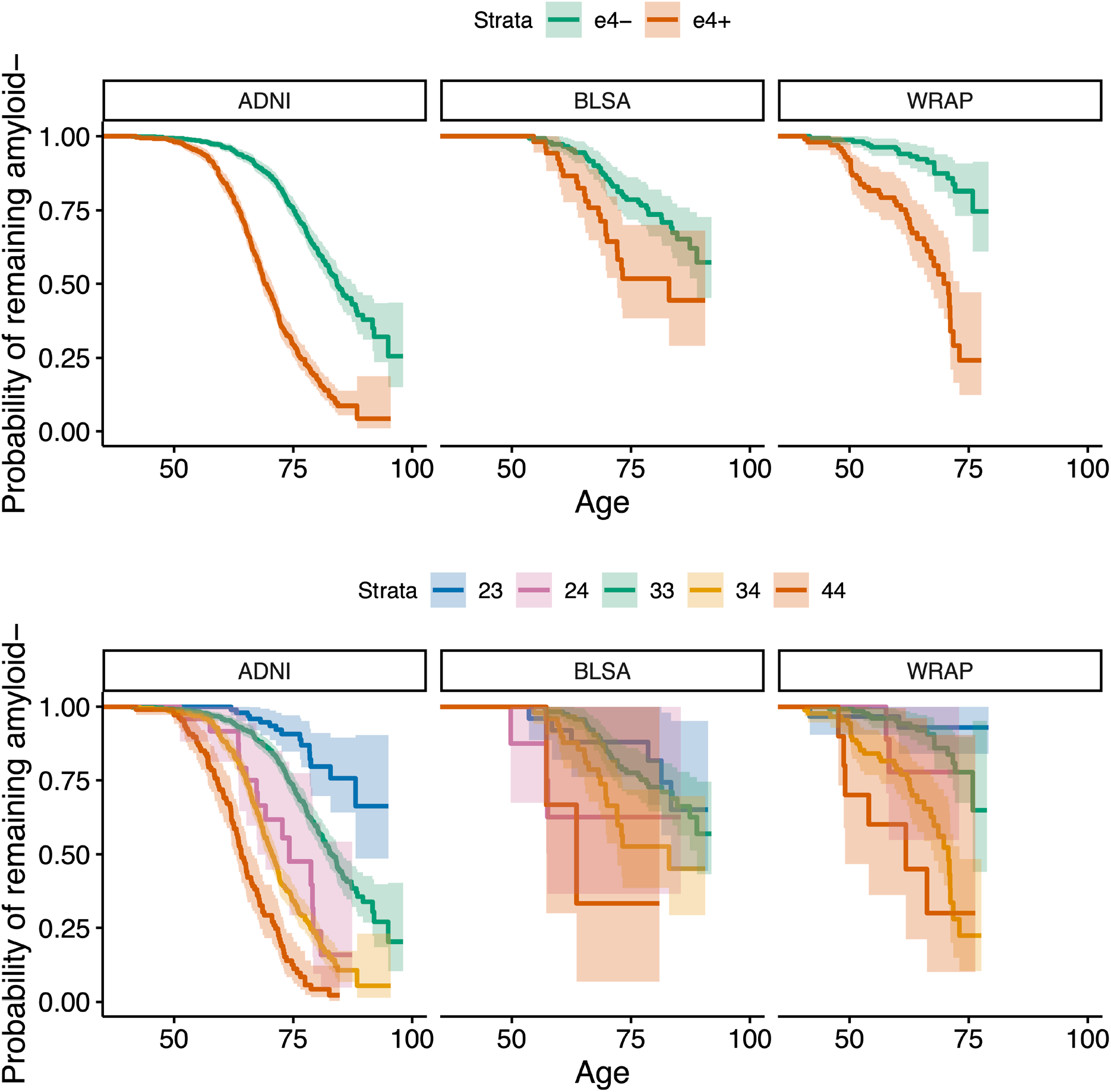
A- survival as a function of age by *APOE-e4* carriage and *APOE* genotype. Kaplan-Meier curves showing A- survival for *APOE* e4 carriers vs. non-carriers (top row) and for individual *APOE* genotypes (bottom row) across all cohorts. Individualized estimates of age A+ (average of the SILA and ODE-GP estimates) were used for analyses. Individuals that were A- at their last available PET scan (i.e., those where Estimate Age A+ < age at last scan) are right censored. Cox proportional hazard models indicated significant differences between e4+ and e4-groups. The hazard for e3e4 and e4e4 groups were higher relative to the e3e3 group in all cohorts, and additionally for the e2e4 group in ADNI. e2e3 carriers had a lower hazard in ADNI.

##### What is the risk of A+ conferred by *APOE*?

Cox proportional hazards models adjusted for sex showed *e4+* status was associated with a greater risk of becoming A+ in each dataset (hazard ratio [HR] (95% CI) in ADNI: HR=3.97 (3.35, 4.70), *P<* 10^−15^; BLSA: HR=2.29 (1.35, 3.90), *P=*0.002; WRAP: HR=5.66 (3.24, 9.86), *P=*10^−9^). Sex differences in A+ risk were not significant and we also did not observe a significant interaction of *e4+* by sex for explaining A+ risk. Results using *APOE* genotype instead of *e4* status for A+ risk are shown in Supplementary Table 8. In ADNI, the *e2e3* genotype was associated with a lower risk of A+ compared to *e3e3* (HR=0.37 (0.22, 0.62), *P=*0.0002), whereas *e2e4* was associated with a higher risk (HR=2.37 (1.40, 4.00), *P=*0.0013). In each data set, *e3e4* and *e4e4* were associated with a higher risk, with the point HR estimate for *e4e4* being higher. The difference in the risk conferred by *e4e4* versus *e3e4* was statistically significant in ADNI only (HR=2.25 (1.78, 2.85), *P<*10^−10^).

##### Does EAOA differ between *APOE* groups?

Accelerated failure time models adjusted for sex indicated *e4*+ individuals had an earlier EAOA (ADNI: *β*=-0.18 (−0.20, −0.16), *P<*10^−15^; BLSA: *β*=-0.14 (−0.23, −0.053), *P=*0.0017; WRAP: *β*=-0.26 (−0.35, −0.17), *P<*10^−8^). Sex differences in EAOA were not significant nor was the interaction between *APOE-e4* status and sex in any dataset. In ADNI, compared to *e3e3*, the *e2e3* group had a later A+ onset (*β*=0.12 (0.064, 0.168), *P=*0.00001), whereas *e2e4* (*β*=-0.11 (−0.18, −0.036), *P=*0.0034), *e3e4* (*β*=-0.14 (−0.16, −0.12), *P<*10^−15^), and *e4e4* (*β*=-0.24 (−0.28, −0.21), *P<*10^−15^) groups had earlier EAOA (Supplementary Table 9). Additionally, the *e4e4* group had an earlier EAOA relative to *e3e4* group (*β*=-0.10 (−0.14, −0.067), *P<*10^−7^). In BLSA, compared to *e3e3*, the *e2e4* (*β*=-0.21 (−0.41, −0.013), *P=*0.037), *e3e4* (*β*=-0.13 (−0.23, −0.035), *P=*0.0076), and *e4e4* (*β*=-0.28 (−0.57, −0.011), *P=*0.059) groups had earlier EAOA. In WRAP, compared to *e3e3*, the *e3e4* (*β*=-0.23 (−0.33, −0.14), *P=*0.000001) and *e4e4* (*β*=-0.35 (−0.53, −0.17), *P=*0.00015) groups had an earlier EAOA.

### Aim 3: EAOA, sex, *APOE* and time to impairment

Participant demographics for this ADNI subset are provided in Table 2. Interactions between sex or *APOE-e4* status with linear and quadratic EAOA terms were not significant (global test *P*=0.44 and *P*=0.17, respectively) in Cox proportional hazard models investigating time from A+ onset to impairment onset. Results for hazard ratios (HR), median unimpaired survival times and ten-year unimpaired survival rates are reported as estimate (95% confidence interval). In the adjusted Cox hazard model *APOE-e4+* had a 77% greater risk of impairment after A+ onset compared to *e4-* (HR=1.77 (1.33, 2.36), *P*<.0001), and males had a 32% lower risk compared to females (HR=0.68, (0.53, 0.88), *P*=.003). Both EAOA and EAOA^2^ terms were statistically significant (each *P*<0.0001). Due to this nonlinearity, we report HRs at multiple EAOA with each HR corresponding to a decade difference in EAOA. Impairment risk was small and not significant for EAOA of 65 vs. 55 (HR=1.06 (0.89, 1.26)), but was significant for EAOA of 75 vs. 65 (63% greater risk; HR=1.63 (1.33, 2.00)), and EAOA of 85 vs. 75 was associated with >2x risk (HR=2.51 (1.71, 3.68)). Groupwise unimpaired survival curves are presented in Figure 5 (panels A, C, D) with median survival times and 10-year unimpaired rates included in supplemental table 12. The median survival time from EAOA to CDR≥1 was 13.57 (12.98-14.08) years with a 10-year post EAOA survival rate of 0.744 (0.703-0.786) across this ADNI subset. Median survival time was shorter for females (12.89 (11.87, 13.66) years) compared to males (14.08 (13.57, 14.55) years), and shorter for *APOE-e4+* (12.73 (11.71, 13.42) years) compared to *e4-* (14.42 (13.97, 15.54) years). The 10-year post EAOA unimpaired survival rate was lower for females (0.699 (0.644-0.755) years) compared to males (0.784 (0.736-0.826) years), and lower for *APOE-e4+* (0.691 (0.637, 0.741) years) vs. *e4-* (0.812 (0.766, 0.857) years). Nonlinear median survival times and 10-year unimpaired survival rates as a function of EAOA are depicted in Figure 5 E-F and supplemental table 10. Analyses using *APOE* genotype (*e3e3, e3e4, e4e4)* instead of *e4+* vs. *e4-* indicated similar results with significant differences between *e3e3* and other genotypes, but not between *e3e4* and *e4e4* genotypes (Panel B of Figure 5 and supplemental results for aim 3).

**FIG 5:**
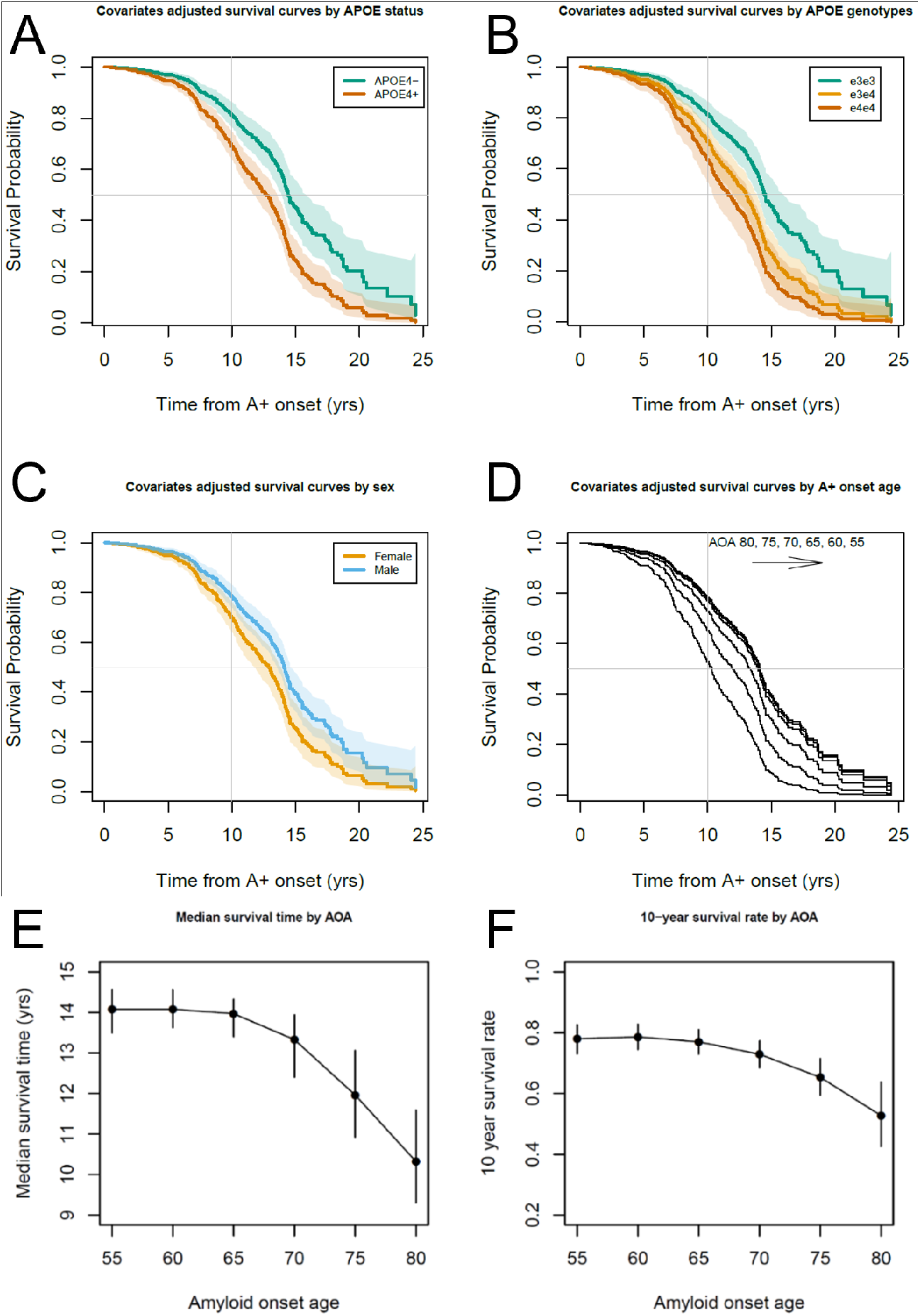
Time from amyloid onset to impairment in ADNI. (**A, B, C, D**) Covariate adjusted survival curves indicating the probability of remaining unimpaired as a function of the time from A+ onset demonstrated that *APOE-e4* carriage, being female, and becoming A+ later in life were all associated with a shorter time from A+ onset to impairment onset (i.e., first CDR≥1). Dotted lines in panels **A, B** and **C** represent 95% confidence intervals of the covariate adjusted survival functions. (**E, F**) Median time to impairment and the 10-year unimpaired rate following A+ onset were nonlinearly associated with the EAOA such that people who became A+ at an older age had greater risk of becoming impaired in a shorter time from A+ onset.

## Discussion

We evaluated three methods to model amyloid accumulation trajectories and estimate A+ onset age from amyloid PET data in three cohorts with different sample characteristics, PET tracers, and quantification methods. These novel methods were then applied to further elucidate the risk and timing of amyloidosis and dementia onset associated with age, sex and *APOE* genotype. Using individualized EAOA, we found that *APOE* genotype, but not sex, significantly impacts EAOA with *e4* homozygotes becoming positive approximately 6 to 10 years earlier than *e3e4* heterozygotes (all cohorts), and approximately two decades earlier than *e3* homozygotes (ADNI). Consistent with prior observations, amyloid accumulation rates are predictable and neither *APOE* genotype, nor sex impacted the rate of amyloid accumulation.^44, 45^ In an ADNI subset, time from A+ onset and eventual cognitive impairment was shorter for *APOE-e4* carriers, females, and persons with higher EAOA. Together, these results provide a major advance in characterizing the temporal course of Alzheimer’s disease on an individual basis and increase understanding of how common risk factors impact disease proteinopathy and clinical progression.

### Method Comparisons and Amyloid PET Inference

Comparisons of the three modeling methods provide insights into model performance, interpretation of amyloid PET binding estimates, and predictability of amyloid aggregation. All methods output EAOA for use in other analyses. A key difference between methods is that ODE-GP and SILA are autonomous whereas GBTM requires more manual steps. For this reason and because we observed similarities between EAOA and amyloid accumulation patterns between methods, we chose to forgo additional SUVR/DVR prediction evaluation for GBTM. The autonomy of ODE-GP and SILA makes these methods efficient for modeling amyloid accumulation in different cohorts and/or subpopulations and easily scalable to applications such as spatiotemporal modeling at the ROI- and voxel-level. Additionally, GBTM was harder to fit in the ADNI dataset, which might suggest ODE-GP and SILA are more robust against differences in PET quantification methods, radiotracers and/or study composition. ODE-GP and SILA also provide non-parametric SUVR/DVR vs. A+ time curves that may better depict the observed nature of amyloid accumulation compared to parametric models that require some assumption of this geometric relationship.

Amyloid accumulation patterns were similar between methods in the A+ range with inter-method differences increasing dramatically below the A+ threshold. This observation, coupled with similarities in DVR/SUVR rates, suggest amyloid accumulation rates are ostensibly consistent between individuals and predictable once underlying amyloid burden sufficiently exceeds the amyloid PET detection limit. Below this threshold, the interpretation of DVR/SUVR gradually transitions from being due to amyloid to being due to stochastic measurement error, which is consistent with PET detection physics, the concept of subthreshold amyloid^46^ and supports previous work proposing two thresholds^47^ to define three interpretation zones: undetectable amyloid, transition, and confidently A+. It follows that EAOA derived from DVR/SUVR values in these different zones will have similar interpretation such that EAOA is more meaningful in the A+ range and becomes less interpretable as reference DVR/SUVR decreases below the A+ threshold. As in our previous work^13^ and a recently published similar work^14^ this ability to predict EAOA in A+ persons enables estimation of A+ disease duration from a single amyloid PET scan, which can be used to gain new insights into how known and unknown factors affect dementia risk and timing in Alzheimer’s disease and may have utility in clinical trials.

### *APOE*, EAOA and Dementia Risk

Our results support literature showing higher *APOE*-*e4* dosage is associated with an earlier age of amyloid and dementia onset.^8, 12, 17-19, 21-23, 45^ Additionally, we observed significant differences in EAOA between almost all compared *APOE* genotypes in ADNI, with similar patterns observed in BLSA and WRAP (smaller sample sizes there likely limited power to detect differences). Individualized EAOA allowed us to show that *e4* carriage shortens the time from A+ onset to dementia onset in a subset of initially unimpaired ADNI participants, even after controlling for age and sex. These findings suggest that *APOE*-*e4* carriers experience a double-hit wherein chronic amyloid accumulation begins earlier in life and the time from amyloid onset to dementia is shorter. Effect sizes for *APOE* genotype differences in A+ onset age far exceeded differences in time from A+ onset to dementia suggesting that *when* amyloid accumulation begins is a key factor explaining *APOE* associated lifetime dementia risk and resilience.

### Clinical Implications

Consistent with our previous reports,^12, 13^ this work suggests individualized EAOA and estimated duration A+ are likely relevant to clinical prognosis and treatment planning. Figures 1 and 3 demonstrate that EAOA varies widely, even within *APOE* genotypes, spanning ages 40 to 90+ years across cohorts. Recruitment differences likely explain some cohort EAOA differences. Nevertheless, the broad range of EAOA highlights the heterogeneity of when amyloid pathology begins to accrue. These results and our previous findings^13^ also underscore the need to consider the magnitude/duration of chronic amyloid exposure derived from quantitative PET imaging to better inform prognostic value compared to binary A+/-. Comparatively less heterogeneity was observed in the time from A+ onset to dementia onset (range 0-25 years post EAOA, Figure 6) in the subset of initially unimpaired A+ ADNI participants. Combining the amyloid chronicity framework^13^ and survival analyses we showed *APOE-e4* carriers, females, and people with older EAOA have shorter time from A+ onset to dementia, and that the effect of EAOA on this timing accelerated rapidly after age 65. In addition to characterizing unimpaired survival times and dementia hazard ratios, these methods can also establish EAOA, sex and *APOE* stratified dementia risk probabilities (e.g., 1-, 5-, 10-year impairment probability) that could inform individualized patient prognoses anchored to EAOA. More work is needed to better understand additional factors that mediate and moderate both the heterogeneity of EAOA and the timing from EAOA to impairment, and to validate risk probabilities in larger and representative samples.

**FIG 6:**
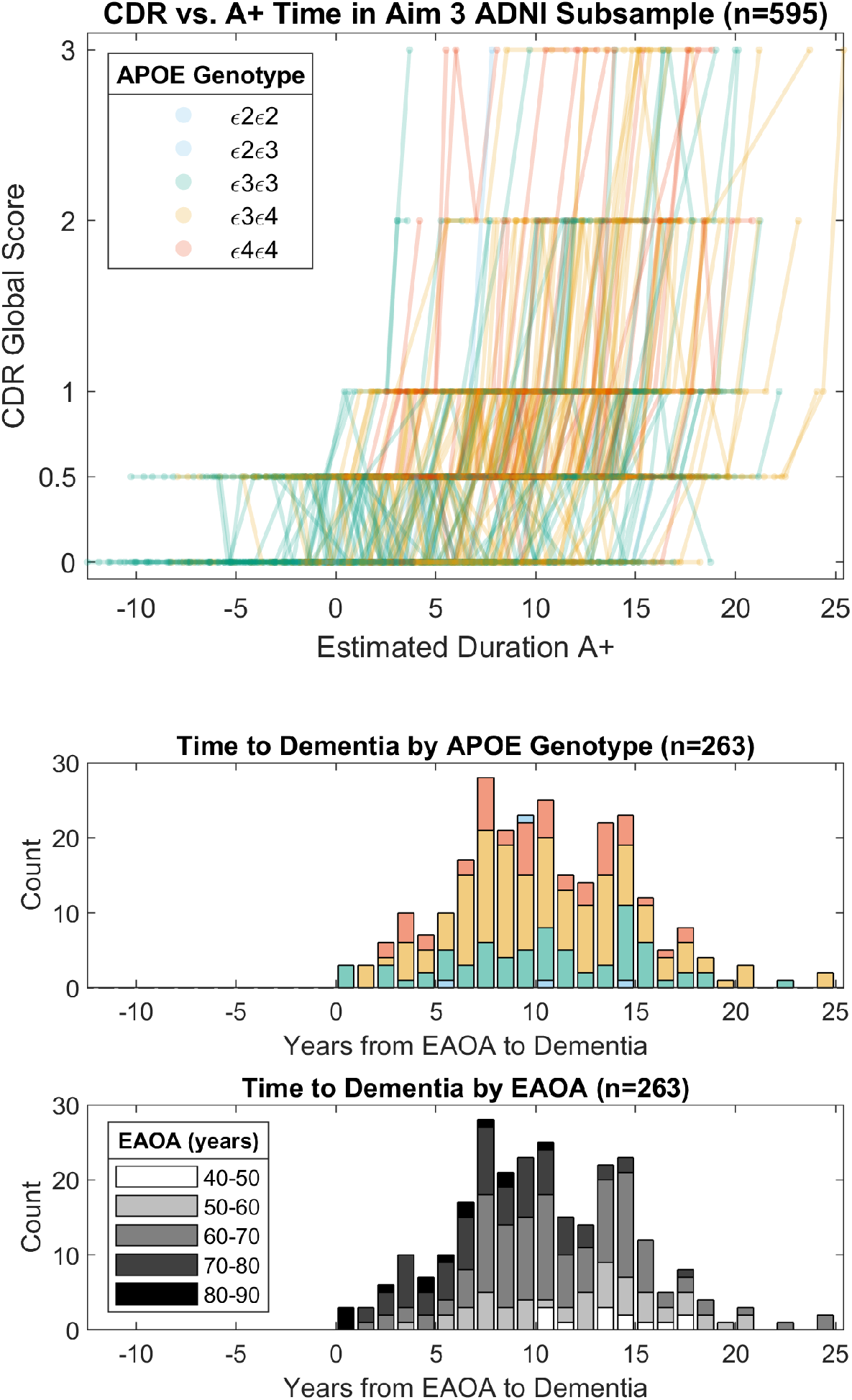
Global CDR scores as a function of the time A+ in ADNI subsample from aim 3 analyses. Observed longitudinal global clinical dementia rating scores are plotted as a function of estimated duration A+ for the ADNI subset used in aim 3 analyses (top). Histograms depict the time from estimated A+ onset age (EAOA) to global CDR = 1 color coded by *APOE* genotype (middle) and by EAOA (bottom) for the subset of participants that became impaired (i.e., global CDR≥1) after A+ onset. Time from A+ onset to global CDR≥1 in this ADNI subsample ranged from 0 to nearly 25 years demonstrating the heterogeneity in the timing of dementia onset relative to A+ onset.

Results regarding the effects of EAOA, sex and *APOE* on the timing between EAOA and dementia have several potential explanations. Similar findings regarding age, but not *APOE* or sex were recently reported using a similar approach.^14^ EAOA shortening the time from A+ onset to impairment may be explained by age-associated increases in comorbidity with other age-related diseases such as vascular disease and other neurodegenerative proteinopathies that co-occur in Alzheimer’s disease.^48^ *APOE-e4* carriage shortening the interval from A+ onset and dementia could represent a pleiotropic susceptibility effect of *APOE* on brain health as *APOE* is also implicated in cerebrovascular health.^49^ Females having shorter time to impairment than males is consistent with but does not fully explain some epidemiologic reports of sex differences in Alzheimer’s disease risk. These results coupled with the observations that sex did not impact EAOA or amyloid accumulation rates is consistent with recent findings that observed no sex differences in Alzheimer’s disease biomarker prevalence but did observe differences in MCI prevalence.^50^ More work is needed to understand the basis for the observed greater vulnerability to dementia for these risk factors. Anchoring such investigations to EAOA may facilitate greater understanding of the relative influence of these and other factors that are associated with increased Alzheimer’s disease risk.

### Strengths and Limitations

This work and other recent publications^3, 7, 11-15, 43, 51^ demonstrate the robustness and utility of the conceptual framework of defining Alzheimer’s disease time using amyloid PET to study disease progression. Strengths of the current study include comparisons and consistency of results across three different methods and cohorts, prediction validation in subsets of observed A- to A+ convertors, and a methodological framework for applying individualized EAOA to characterize biomarker and dementia timing and risk in Alzheimer’s disease. Further work is needed to better understand the impact of PET quantification methods, radiotracers, different PET and MRI scanners, and cohort composition on temporal amyloid models, and the extent to which EAOA can be broadly harmonized. Additionally, this study used convenience samples which are skewed toward highly educated, white persons. More work is needed to collect biomarkers in diverse populations and to test the extent these models and concepts are applicable outside of the participants studied here.

## Conclusions

This work demonstrates multiple methods for modeling amyloid accumulation and estimating the onset age and duration of amyloid positivity from PET imaging. These novel approaches and outcomes can be used to more directly investigate the timing of cognitive and pathological events in Alzheimer’s disease on an individual basis, and better understand factors that may modify this timing.

## Data Availability

Data from the WRAP (https://wrap.wisc.edu) and BLSA (https://blsa.nih.gov) can be requested through an online submission process. ADNI data can be obtained via the ADNI and LONI websites (http://adni.loni.usc.edu/).

https://blsa.nih.gov

http://adni.loni.usc.edu/

## Abbreviations

A+/-: elevated or non-elevated amyloid PET
ADNI: Alzheimer’s Disease Neuroimaging Initiative
APOE: Apolipoprotein E
BLSA: Baltimore Longitudinal Study of Aging
CDR: clinical dementia rating
DVR: distribution volume ratio
EAOA: estimated amyloid onset age
GBTM: Group-Based Trajectory Modeling
MCI: mild cognitive impairment
ODE-GP: Ordinary Differential Equation-Gaussian Process
PAC: Preclinical AD Consortium
PiB: [^11^C]Pittsburgh compound B
SILA: Sampled Iterative Local Approximation
SUVR: standard uptake value ratio
WRAP: Wisconsin Registry for Alzheimer’s Prevention

## Acknowledgements

We would like to thank all participants and their families and the many study teams that make this work possible. This work was supported in part by the Intramural Research Program of the National Institute on Aging, National Institutes of Health. Data collection and sharing for this project was funded by the Alzheimer’s Disease Neuroimaging Initiative (ADNI) (National Institutes of Health Grant U01 AG024904) and DOD ADNI (Department of Defense award number W81XWH-12-2-0012). ADNI is funded by the National Institute on Aging, the National Institute of Biomedical Imaging and Bioengineering, and through generous contributions from the following: AbbVie, Alzheimer’s Association; Alzheimer’s Drug Discovery Foundation; Araclon Biotech; BioClinica, Inc.; Biogen; Bristol-Myers Squibb Company; CereSpir, Inc.; Cogstate; Eisai Inc.; Elan Pharmaceuticals, Inc.; Eli Lilly and Company; EuroImmun; F. Hoffmann-La Roche Ltd and its affiliated company Genentech, Inc.; Fujirebio; GE Healthcare; IXICO Ltd.; Janssen Alzheimer Immunotherapy Research & Development, LLC.; Johnson & Johnson Pharmaceutical Research & Development LLC.; Lumosity; Lundbeck; Merck & Co., Inc.; Meso Scale Diagnostics, LLC.; NeuroRx Research; Neurotrack Technologies; Novartis Pharmaceuticals Corporation; Pfizer Inc.; Piramal Imaging; Servier; Takeda Pharmaceutical Company; and Transition Therapeutics. The Canadian Institutes of Health Research is providing funds to support ADNI clinical sites in Canada. Private sector contributions are facilitated by the Foundation for the National Institutes of Health (www.fnih.org). The grantee organization is the Northern California Institute for Research and Education, and the study is coordinated by the Alzheimer’s Therapeutic Research Institute at the University of Southern California. ADNI data are disseminated by the Laboratory for Neuro Imaging at the University of Southern California.

## Supplementary material

Supplementary material is available at *Brain* online.

