## Supplement for "Multi-method investigation of factors influencing amyloid onset and impairment in three cohorts"

### Supplemental Material

#### Supplemental Methods

##### Amyloid Trajectory Modeling Methods

###### Group-Based Trajectory Modeling (GBTM)

Group-based trajectory modeling (GBTM) is an extension of the initial WRAP amyloid chronicity publication.[1] In this approach, we used GBTM[2, 3] to identify functions that characterize age-related amyloid DVR or SUVR accumulation patterns in each cohort. For the WRAP sample, we used the previously published functions.[1] For ADNI and BLSA, we followed the same iterative GBTM process first to identify the optimal number of groups, and subsequently, to identify the shapes of the age-related accumulation functions. The final set of functions was selected for each cohort based on model fits and "common sense" criteria (e.g., functions could not overlap with each other in any portion of the observed age range of the development set). After the set of models was selected for each cohort, each function in the set was solved to determine the age at which the DVR or SUVR was equal to the A+ cut off for the "trajectory group" represented by that function. Using each person's most recent scan, the residuals of the observed DVR or SUVR were calculated for each GBTM function and the residuals were then used to estimate the probability of membership in each of the trajectory groups. In the original publication on this method, we used the residuals from all four of the GBTM functions and applied Bayes' theorem to estimate the probability of group membership. In this paper, we simplified the approach to use the residuals of the two closest GBTM functions to estimate probability of group membership for the two closest groups. These probabilities were then used as weights as described in Koscik, Betthauser, et al to estimate age of A+ onset. To obtain A+ duration (termed A+ chronicity in Koscik, et al.) for a measurement of interest (e.g., A+ duration at baseline cognitive assessment or A+ duration at amyloid scan), we subtracted estimated age of A+ onset from age of measurement such that positive numbers indicate estimated number of years above the A+ cut-off. We applied piecewise regression techniques to the chronicity-aligned data from each cohort and developed a set of equations characterizing the accumulation trajectory. These equations were then used to estimate amyloid levels as well as time between different amyloid levels.

###### Ordinary Differential Equation – Gaussian Process

The Ordinary Differential Equation – Gaussian Process (ODE-GP) algorithm aims at fitting a non-parametric ordinary differential equation (ODE) to data. In this work, the data are the SUVR or DVR values recorded longitudinally for the participants in each study (WRAP, BLSA, and ADNI). Fitting parametric ODEs or their discrete versions to data has a long history and goes by other names, including state space methods, dynamical systems identification, and probabilistic graphical models[4, 5] , and is very common in engineering and control. The non-parametric setting has only been studied recently[6-9] thanks to progresses in nonparametric regression, both from the statistical frequentist point of view, using the theory of Reproducing Kernel Hilbert space (RKHSs)[10] and from the Bayesian point of view, using Gaussian Processes (GPs)[11].

The ODE-GP algorithm is a gradient matching method, see [8], which proceeds in two steps. The first step consists in estimating gradients, or longitudinal slopes of SUVR or DVR, one for each subject, using standard simple linear regression. The second step consists in solving a non-standard GP regression problem using the data from all subjects in a cohort, considering both the noise in the measurements and the noise in the estimated gradients. An approximate solution is provided using a Taylor expansion at order two of the Gaussian process. We choose a standard kernel: the Gaussian kernel. The radius of the kernel is estimated by maximizing the marginal likelihood of the data. The fitted model allows for the prediction of past and future DVR or SUVR values for a subject in each cohort using a single DVR or SUVR value at a given age providing an initial condition for the ODE. The prediction is made by integrating numerically the ODE using the Euler method. Since the fitted model is autonomous (time-invariant), we summarize the fitted data with a growth curve defining the DVR or SUVR as function of the time since a given DVR or SUVR level is achieved.

**Sampled Iterative Local Approximation**

Sampled iterative local approximation (SILA) uses a three-step process that includes discrete sampling of amyloid accumulation rates, smoothing and numerical integration to generate a nonparametric amyloid vs. time curve. Individual estimates of duration A+ and age A+ are estimated by solving the nonparametric curve for the duration A+ corresponding to a given DVR or SUVR value at one or more PET observations. The first step in the modeling algorithm applies discrete sampling to within-person annualized longitudinal DVR or SUVR slopes to establish the 'observed' relationship between the annualized amyloid accumulation rate and the level of amyloid. Query DVR or SUVR values are established by dividing the full range of observed values into 150 equally spaced values. For each query value, the mean SUVR or DVR rate is calculated for participants whose observations intersect the query value. The second step of the algorithm optimizes a robust weighted local 2$^{nd}$ degree polynomial smoothing kernel (rloess option in MATLAB smooth function) applied to the numerical rate vs level data by minimizing the weighted sum of squared backward prediction residuals (i.e. predicting a first DVR or SUVR value using the last observation as a reference). A weighting function is applied for optimizing the kernel size that de-weights the sum of squares for A- cases by multiplying them by the ratio of A+ to A- cases. This prioritizes A+ cases for the modeled amyloid vs. time function. The third step uses Euler's method to numerically integrate the smoothed amyloid rate vs. amyloid level function to produce an amyloid level vs. time curve. For this study, a 0.25 year step size was used for numerical integration with the initial condition that the DVR or SUVR positivity threshold corresponds to time = 0 years. Thus, the time axis corresponds to the number or years a person is A+. The algorithm terminates if either the number of observations for a given amyloid level was less than two, the average slope was ≤0, or the max iteration limit of 200 (corresponding to 50 years) was observed. A+ duration was estimated for individual participants at a reference observation (single scan) by numerically solving the nonparametric amyloid vs A+ duration function for time inputting the observed DVR or SUVR value at the reference scan. Age of A+ was then estimated by subtracting the estimated duration A+ from the age at the reference scan. The algorithm also includes a within-person least squares option to solve for time A+ using multiple observations, but this was not used in the current study. Linear extrapolation of the first or last three years of the modeled curve was used to estimate age and duration A+ in reference values were out of range of the modeled nonparametric curve. The duration A+ at reference scan for cases whose observations were below the modeled DVR or SUVR range was truncated such that the reference scan for each participant was aligned to the earliest modeled point on the nonparametric amyloid vs. time curve. Forward and backward DVR or SUVR prediction was accomplished by solving the amyloid vs. time curve for DVR or SUVR corresponding to the duration of A+ at the time of the reference scan minus the time of the target scan being predicted.

#### Generation of model prediction training and testing sets

ADNI's sample size allowed us to use holdout validation to test model prediction in a larger test sample that included people with multiple scans. Specifically, we established age x SUVR strata (baseline age decades, beginning with <65; SUVR A+ and -) and randomly assigned people within strata to one of three partitions. Two partitions were combined to form the training set and the third partition along with any single scan participant data was used as the testing set. The folds for the 10-fold cross-validation studies were created by randomly assigning each participant to one of the 10 folds and ensuring that the resulting folds had similar distributions for age and DVR or SUVR values.

### Supplemental Results

#### Aim 3: Time from EAOA to impairment onset by *APOE* genotype

In a separate adjusted Cox proportional hazards model, we investigated *APOE* genotype rather than dichotomous *e4* carriage. In reference to the *e3e3* group, the *e3e4* group had a 67% greater risk (HR=1.67 (1.24, 2.26), *P*=.0008) and the *e4e4* group had over two-fold higher risk (HR=2.20 (1.50, 3.23), *P*<.0001), but the relative risk conferred by *e4e4* vs. *e3e4* was not statistically significant (HR=1.31, (0.95, 1.83), *P*=0.104). Covariate adjusted survival curves for each *APOE* genotype are shown in Figure 5B with median survival time and 10-year unimpaired rate summarized in supplemental table 13. Median survival time from EAOA to impairment onset was lowest for *e4e4* carriers (11.61 (10.35-13.22) years), followed by *e3e4* carriers (13.03 (11.96-13.66) years) and then *e3e3* carriers (14.46 (13.87-15.80) years). The unimpaired survival rate ten years after EAOA was similarly lowest for *e4e4* carriers (0.635 (0.540-0.728)) followed by *e3e4* carriers (0.708 (0.649-0.763)) and then *e3e3* carriers (0.814 (0.763-0.859)).

### SUPPLEMENTAL Tables

##### Overview of Supplemental Tables and Figures

The supplemental tables and figures give additional information about amyloid positivity thresholds, and analyses evaluating model performance, amyloid accumulation rates, amyloid onset age and time from amyloid positivity to impairment onset. Supplemental table 1 provides details regarding how amyloid PET scans were quantified and how amyloid positivity thresholds were determined for each cohort. Supplemental tables 2-9 and supplemental figures 1-7 provide details regarding analyses for aim one investigating model performance. Supplemental figures 8-9 give additional information regarding rates of amyloid accumulation as a function of amyloid level by APOE *e4* carriage and sex, respectively. Supplemental tables 10-11 provide additional information for aim two analyses investigating the risk of amyloid positivity and accelerate failure time model outcomes by *APOE* genotype. Supplemental tables 12-13 provide median survival times and 10-year unimpaired rates for analyses in aim three investigating factors associated with time from A+ onset to impairment onset.

**Supplemental Table 1: Amyloid positivity thresholds and methods for each cohort**

| Cohort | A+ Threshold | Threshold Derivation Method |
|---|---|---|
| ADNI | >0.8 $SUVR_{WM}$ | ROC of BAI processed data with Berkeley stratified A+/- |
| BLSA | >1.066 $DVR_{SRTM-LRSC}$ | Two-class Gaussian mixture model |
| WRAP | >1.19 $DVR_{LGA}$ | ROC with visual rating of A+/- as the standard for comparison |

$SUVR_{WM}$ = standard uptake value ratio using the composite white matter as reference region from the Banner Alzheimer's Institute processed data; $DVR_{SRTM-LRSC}$ = distribution volume ratio estimated using cerebellar gray matter reference region and the simplified reference tissue method with spatial constraint; $DVR_{LGA}$ = distribution volume ratio estimated using cerebellar gray matter reference region and Logan graphical analysis; ROC = receiver operating characteristic

**Supplemental Table 2: Inter-method EAOA correlations for A+ participants**

| Cohort | Contrast | n | $r_P$ (95% CI) | $p_P$ |
|---|---|---|---|---|
| ADNI | ODE-GP v. SILA | 614 | 1.00 (1.00, 1.00) | <0.001 |
| ADNI | ODE-GP v. GBTM | 614 | 0.91 (0.89, 0.92) | <0.001 |
| ADNI | GBTM v. SILA | 614 | 0.88 (0.86, 0.89) | <0.001 |
| BLSA | ODE-GP v. SILA | 65 | 0.99 (0.98, 0.99) | <0.001 |
| BLSA | ODE-GP v. GBTM | 65 | 0.92 (0.87, 0.95) | <0.001 |
| BLSA | GBTM v. SILA | 65 | 0.88 (0.81, 0.93) | <0.001 |
| WRAP | ODE-GP v. SILA | 62 | 1.00 (1.00, 1.00) | <0.001 |
| WRAP | ODE-GP v. GBTM | 62 | 0.98 (0.97, 0.99) | <0.001 |
| WRAP | GBTM v. SILA | 62 | 0.99 (0.98, 0.99) | <0.001 |

Models trained on all participants with two or more amyloid PET scans. EAOA and A+ status were determined using each participant's last available amyloid PET observation. EAOA = estimated age of A+ onset; n = the number of A+ participants; $r_P$ = Pearson correlation coefficient; $p_P$ = p-value for Pearson correlation

**Supplemental Table 3: Inter-method EAOA correlations for A- participants**

| Cohort | Contrast | n | $r_P$ (95% CI) | $p_P$ |
|---|---|---|---|---|
| ADNI | ODE-GP v. SILA | 545 | 0.95 (0.94, 0.96) | <0.001 |
| ADNI | ODE-GP v. GBTM | 545 | 0.85 (0.82, 0.87) | <0.001 |
| ADNI | GBTM v. SILA | 601 | 0.77 (0.74, 0.80) | <0.001 |
| BLSA | ODE-GP v. SILA | 73 | 0.90 (0.84, 0.93) | <0.001 |
| BLSA | ODE-GP v. GBTM | 73 | 0.80 (0.70, 0.87) | <0.001 |
| BLSA | GBTM v. SILA | 142 | 0.76 (0.68, 0.82) | <0.001 |
| WRAP | ODE-GP v. SILA | 78 | 0.76 (0.64, 0.84) | <0.001 |
| WRAP | ODE-GP v. GBTM | 78 | 0.89 (0.83, 0.93) | <0.001 |
| WRAP | GBTM v. SILA | 210 | 0.79 (0.73, 0.84) | <0.001 |

Models trained on all participants with two or more amyloid PET scans. Comparisons of SILA and GBTM with ODE-GP are limited to participants where ODE-GP produces a finite EAOA. EAOA = estimated age of A+ onset; n = the number of A- participants with finite EAOA; $r_P$ = Pearson correlation coefficient; $p_P$ = p-value for Pearson correlation

**Supplemental Table 4: EAOA accuracy and midpoint error in A- to A+ convertors**

| Cohort | ADNI | BLSA | WRAP |
|---|---|---|---|
| Number of A- to A+ convertors in sample | 72 | 6 | 22 |
| GBTM Accuracy, % (CI) [$n_{corr}$] | 25.0%, (15.0, 35.0) [18] | 16.7%, (-13.2, 46.5), [1] | 63.6%, (43.5, 83.7), [14] |
| ODE-GP Accuracy, % (CI) [$n_{corr}$] | 50.0%, (38.5, 61.6) [36] | 66.7%, (29.0, 104.4), [4] | 54.6%, (33.7, 75.4), [12] |
| SILA Accuracy, % (CI) [$n_{corr}$] | 61.1%, (49.9, 72.4) [44] | 33.3%, (-4.4, 71.1), [2] | 77.3%, (59.8, 94.8), [17] |
| GBTM conversion midpoint error, mean (CI) | -3.65 (-4.55, -2.75) | 1.96 (0.50, 3.42) | -0.05 (-0.98, 0.89) |
| ODE-GP conversion midpoint error, mean (CI) | -1.29 (-1.80, -0.79) | -0.64 (-1.36, 0.08) | -1.50 (-2.38, -0.62) |
| SILA conversion midpoint error mean, (CI) | -0.42 (-0.83, -0.01) | 1.10 (0.53, 1.68) | -0.64 (-1.35, 0.08) |

Accuracy is defined as the number of times age at last A- scan ≤ EAOA ≤ age at first A+ scan divided by the total number of participants within each sample that converted from A- to A+ and remained A+ during the remainder of the study. Conversion midpoint error is the difference between EAOA and the midpoint age between last A- and first A+ scans. CI represents the 95% confidence interval. $n_{corr}$ represents the number of times EAOA fell between the last A- and first A+ scan.

**Supplemental Table 5: Out-of-sample prediction summary statistics**

| Method | Direction | SSQ | SSQ A+ | SSQ A- | RMSE | MAE | Balanced Accuracy | Sensitivity | Specificity |
|---|---|---|---|---|---|---|---|---|---|
| | | | | ADNI Holdout Cross-validation | | | | | |
| ODE-GP | backward | 7.79 | 3.84 | 3.95 | 0.032 | 0.025 | 0.98 | 0.99 | 0.97 |
| SILA | backward | 8.68 | 4.32 | 4.36 | 0.035 | 0.027 | 0.95 | 0.92 | 0.98 |
| GBTM | backward | 12.14 | 4.53 | 7.61 | 0.049 | 0.040 | 0.95 | 0.97 | 0.94 |
| ODE-GP | forward | 10.59 | 5.80 | 4.80 | 0.043 | 0.032 | 0.97 | 0.98 | 0.95 |
| SILA | forward | 12.71 | 6.62 | 6.09 | 0.051 | 0.036 | 0.94 | 1.00 | 0.89 |
| GBTM | forward | 11.75 | 5.44 | 6.31 | 0.048 | 0.032 | 0.98 | 0.97 | 0.98 |
| | | | | ADNI 10-Fold Cross-validation | | | | | |
| ODE-GP | backward | 23.78 | 12.07 | 11.70 | 0.032 | 0.025 | 0.97 | 0.98 | 0.96 |
| SILA | backward | 25.35 | 13.01 | 12.33 | 0.034 | 0.027 | 0.94 | 0.91 | 0.97 |
| ODE-GP | forward | 32.57 | 19.63 | 12.94 | 0.044 | 0.033 | 0.96 | 0.94 | 0.97 |
| SILA | forward | 36.73 | 21.14 | 15.59 | 0.050 | 0.037 | 0.94 | 0.98 | 0.91 |
| | | | | BLSA 10-Fold Cross-validation | | | | | |
| ODE-GP | backward | 3.25 | 1.72 | 1.53 | 0.023 | 0.015 | 0.99 | 0.98 | 1.00 |
| SILA | backward | 3.12 | 1.58 | 1.54 | 0.022 | 0.013 | 0.99 | 0.98 | 1.00 |
| ODE-GP | forward | 5.02 | 2.08 | 2.94 | 0.035 | 0.025 | 0.91 | 0.91 | 0.91 |
| SILA | forward | 4.65 | 1.83 | 2.82 | 0.033 | 0.016 | 0.92 | 0.93 | 0.91 |
| | | | | WRAP 10-Fold Cross-validation | | | | | |
| ODE-GP | backward | 5.82 | 1.29 | 4.53 | 0.033 | 0.025 | 0.95 | 0.96 | 0.95 |
| SILA | backward | 6.37 | 1.37 | 5.00 | 0.036 | 0.028 | 0.95 | 0.91 | 0.99 |
| ODE-GP | forward | 11.69 | 3.28 | 8.41 | 0.065 | 0.051 | 0.83 | 0.75 | 0.92 |
| SILA | forward | 10.42 | 2.97 | 7.45 | 0.058 | 0.037 | 0.82 | 0.77 | 0.86 |

Forward prediction estimated DVR/SUVR at the last scan using the first scan as a reference to estimate age A+. Backward prediction estimated DVR/SUVR at the first scan using the last scan as a reference to estimate age A+. SSQ A+ and SSQ A- represent the components of the sum of squared DVR/SUVR residuals for amyloid positive and amyloid negative participants, respectively. Balanced accuracy adjusts for the uneven proportions of A+ and A- cases in each cohort.

ODE-GP = ordinary differential equation-Gaussian process, SILA = Sampled iterative local approximation, GBTM = group-based trajectory modeling, SSQ = sum of squares, A+ = amyloid positive, A- = amyloid negative, RMSE = root mean squared error, MAE = median absolute error ADNI = Alzheimer's Disease Neuroimaging Initiative, BLSA = Baltimore Longitudinal Study on Aging, WRAP = Wisconsin Registry for Alzheimer's Prevention, ODE-GP = ordinary differential equation-Gaussian process, SILA = Sampled iterative local approximation, GBTM = group-based trajectory modeling

**Supplemental Table 6: Spearman Correlations with Prediction Residuals for ADNI, Holdout cross-validation**

| Direction | Method | Predictor | $r_s$ (95% CI) | $r_s$ (95% CI), A+ | $r_s$ (95% CI), A- |
|---|---|---|---|---|---|
| Backwards | GBTM | Ref SUVR | 0.45 (0.33, 0.58) | 0.08 (-0.04, 0.21) | 0.57 (0.44, 0.69) |
| Backwards | GBTM | Time to Ref | 0.26 (0.14, 0.39) | 0.46 (0.33, 0.58) | 0.02 (-0.10, 0.15) |
| Backwards | GBTM | Age at Ref | -0.21 (-0.33, -0.08) | -0.34 (-0.46, -0.21) | -0.28 (-0.40, -0.15) |
| Backwards | ODE-GP | Ref SUVR | -0.01 (-0.14, 0.11) | 0.13 (0.01, 0.26) | -0.30 (-0.43, -0.17) |
| Backwards | ODE-GP | Time to Ref | -0.03 (-0.15, 0.10) | 0.17 (0.04, 0.29) | -0.22 (-0.34, -0.09) |
| Backwards | ODE-GP | Age at Ref | 0.04 (-0.09, 0.16) | 0.19 (0.07, 0.32) | -0.13 (-0.25, 0.00) |
| Backwards | SILA | Ref SUVR | 0.06 (-0.07, 0.19) | 0.15 (0.02, 0.27) | -0.21 (-0.33, -0.08) |
| Backwards | SILA | Time to Ref | -0.15 (-0.27, -0.02) | 0.03 (-0.10, 0.15) | -0.35 (-0.47, -0.22) |
| Backwards | SILA | Age at Ref | 0.04 (-0.08, 0.17) | 0.21 (0.08, 0.33) | -0.14 (-0.27, -0.02) |
| Forwards | GBTM | Ref SUVR | 0.61 (0.49, 0.74) | 0.22 (0.10, 0.35) | 0.69 (0.56, 0.81) |
| Forwards | GBTM | Time to Ref | -0.11 (-0.23, 0.02) | 0.01 (-0.11, 0.14) | -0.04 (-0.16, 0.09) |
| Forwards | GBTM | Age at Ref | -0.28 (-0.41, -0.16) | -0.57 (-0.69, -0.44) | -0.22 (-0.35, -0.09) |
| Forwards | ODE-GP | Ref SUVR | -0.19 (-0.31, -0.06) | -0.35 (-0.47, -0.22) | -0.03 (-0.16, 0.09) |
| Forwards | ODE-GP | Time to Ref | -0.04 (-0.16, 0.09) | 0.13 (0.00, 0.25) | -0.19 (-0.31, -0.06) |
| Forwards | ODE-GP | Age at Ref | -0.04 (-0.16, 0.09) | -0.19 (-0.32, -0.06) | 0.10 (-0.03, 0.22) |
| Forwards | SILA | Ref SUVR | -0.23 (-0.36, -0.11) | -0.36 (-0.48, -0.23) | -0.11 (-0.24, 0.01) |
| Forwards | SILA | Time to Ref | -0.21 (-0.34, -0.09) | -0.06 (-0.18, 0.07) | -0.40 (-0.52, -0.27) |
| Forwards | SILA | Age at Ref | -0.08 (-0.21, 0.05) | -0.22 (-0.35, -0.10) | 0.05 (-0.08, 0.17) |

**Supplemental Table 7: Spearman Correlations with Prediction Residuals for ADNI, 10-fold cross-validation**

| Direction | Method | Predictor | $r_s$ (95% CI) | $r_s$ (95% CI), A+ | $r_s$ (95% CI), A- |
|---|---|---|---|---|---|
| Backwards | ODE-GP | Ref SUVR | -0.05 (-0.12, 0.02) | 0.04 (-0.03, 0.11) | -0.23 (-0.30, -0.16) |
| Backwards | ODE-GP | Time to Ref | 0.00 (-0.07, 0.07) | 0.14 (0.06, 0.21) | -0.15 (-0.23, -0.08) |
| Backwards | ODE-GP | Age at Ref | -0.01 (-0.08, 0.07) | 0.12 (0.04, 0.19) | -0.15 (-0.22, -0.08) |
| Backwards | SILA | Ref SUVR | 0.01 (-0.06, 0.09) | 0.02 (-0.05, 0.09) | -0.16 (-0.23, -0.09) |
| Backwards | SILA | Time to Ref | -0.12 (-0.19, -0.05) | -0.01 (-0.08, 0.07) | -0.27 (-0.34, -0.20) |
| Backwards | SILA | Age at Ref | 0.00 (-0.07, 0.07) | 0.11 (0.03, 0.18) | -0.15 (-0.22, -0.07) |
| Forwards | ODE-GP | Ref SUVR | -0.15 (-0.22, -0.08) | -0.21 (-0.28, -0.14) | -0.05 (-0.12, 0.03) |
| Forwards | ODE-GP | Time to Ref | 0.00 (-0.07, 0.08) | 0.05 (-0.02, 0.12) | -0.08 (-0.15, -0.01) |
| Forwards | ODE-GP | Age at Ref | 0.01 (-0.06, 0.09) | -0.08 (-0.16, -0.01) | 0.12 (0.05, 0.20) |
| Forwards | SILA | Ref SUVR | -0.19 (-0.27, -0.12) | -0.20 (-0.28, -0.13) | -0.16 (-0.23, -0.08) |
| Forwards | SILA | Time to Ref | -0.18 (-0.25, -0.10) | -0.13 (-0.20, -0.06) | -0.29 (-0.36, -0.22) |
| Forwards | SILA | Age at Ref | -0.03 (-0.10, 0.04) | -0.11 (-0.18, -0.04) | 0.06 (-0.01, 0.14) |

**Supplemental Table 8: Spearman Correlations with Prediction Residuals for BLSA, 10-fold cross-validation**

| Direction | Method | Predictor | $r_s$ (95% CI) | $r_s$ (95% CI), A+ | $r_s$ (95% CI), A- |
|---|---|---|---|---|---|
| Backwards | ODE-GP | Ref DVR | -0.03 (-0.19, 0.14) | 0.12 (-0.05, 0.28) | 0.20 (0.04, 0.37) |
| Backwards | ODE-GP | Time to Ref | 0.04 (-0.12, 0.21) | 0.18 (0.01, 0.34) | -0.02 (-0.19, 0.14) |
| Backwards | ODE-GP | Age at Ref | -0.09 (-0.26, 0.08) | 0.03 (-0.14, 0.20) | -0.13 (-0.30, 0.04) |
| Backwards | SILA | Ref DVR | -0.06 (-0.22, 0.11) | 0.23 (0.07, 0.40) | -0.18 (-0.35, -0.02) |
| Backwards | SILA | Time to Ref | -0.01 (-0.18, 0.15) | 0.03 (-0.14, 0.20) | -0.05 (-0.22, 0.11) |
| Backwards | SILA | Age at Ref | 0.02 (-0.15, 0.19) | 0.06 (-0.11, 0.22) | -0.02 (-0.18, 0.15) |
| Forwards | ODE-GP | Ref DVR | -0.30 (-0.47, -0.13) | -0.43 (-0.60, -0.27) | -0.63 (-0.80, -0.47) |
| Forwards | ODE-GP | Time to Ref | 0.05 (-0.12, 0.22) | 0.07 (-0.10, 0.23) | 0.03 (-0.14, 0.20) |
| Forwards | ODE-GP | Age at Ref | 0.09 (-0.08, 0.26) | 0.03 (-0.13, 0.20) | 0.13 (-0.04, 0.29) |
| Forwards | SILA | Ref DVR | -0.28 (-0.45, -0.12) | -0.34 (-0.51, -0.18) | -0.44 (-0.60, -0.27) |
| Forwards | SILA | Time to Ref | -0.04 (-0.21, 0.12) | -0.03 (-0.19, 0.14) | -0.05 (-0.21, 0.12) |
| Forwards | SILA | Age at Ref | 0.00 (-0.16, 0.17) | -0.01 (-0.17, 0.16) | 0.04 (-0.13, 0.20) |

**Supplemental Table 9: Spearman Correlations with Prediction Residuals for WRAP, 10-fold cross-validation**

| Direction | Method | Predictor | $r_s$ (95% CI) | $r_s$ (95% CI), A+ | $r_s$ (95% CI), A- |
|---|---|---|---|---|---|
| Backwards | ODE-GP | Ref DVR | -0.18 (-0.33, -0.03) | 0.29 (0.14, 0.44) | 0.09 (-0.06, 0.24) |
| Backwards | ODE-GP | Time to Ref | 0.05 (-0.10, 0.19) | -0.02 (-0.16, 0.13) | 0.04 (-0.11, 0.19) |
| Backwards | ODE-GP | Age at Ref | -0.06 (-0.21, 0.09) | -0.19 (-0.34, -0.05) | 0.07 (-0.08, 0.22) |
| Backwards | SILA | Ref DVR | -0.41 (-0.56, -0.26) | 0.20 (0.05, 0.35) | -0.43 (-0.58, -0.28) |
| Backwards | SILA | Time to Ref | -0.04 (-0.19, 0.11) | -0.04 (-0.19, 0.11) | -0.08 (-0.23, 0.07) |
| Backwards | SILA | Age at Ref | 0.02 (-0.13, 0.16) | -0.05 (-0.20, 0.10) | 0.13 (-0.02, 0.28) |
| Forwards | ODE-GP | Ref DVR | -0.36 (-0.51, -0.21) | -0.01 (-0.16, 0.14) | -0.47 (-0.61, -0.32) |
| Forwards | ODE-GP | Time to Ref | 0.06 (-0.09, 0.20) | 0.04 (-0.11, 0.19) | 0.07 (-0.08, 0.21) |
| Forwards | ODE-GP | Age at Ref | 0.03 (-0.11, 0.18) | 0.43 (0.29, 0.58) | 0.01 (-0.14, 0.16) |
| Forwards | SILA | Ref DVR | -0.26 (-0.41, -0.11) | -0.02 (-0.17, 0.13) | -0.36 (-0.51, -0.21) |
| Forwards | SILA | Time to Ref | -0.04 (-0.19, 0.11) | 0.02 (-0.13, 0.17) | -0.04 (-0.19, 0.10) |
| Forwards | SILA | Age at Ref | 0.03 (-0.12, 0.17) | 0.20 (0.05, 0.35) | 0.00 (-0.15, 0.15) |

**Supplementary Table 10: Hazard ratios for risk of amyloid positivity by *APOE* genotype**

| | Data set | | |
|---|---|---|---|
| ***APOE* genotype** | *ADNI* | *BLSA* | *WRAP* |
| ***e2e3*** | 0.37 (0.22, 0.62)*** | 0.89 (0.37, 2.12) | 0.38 (0.09, 1.68) |
| ***e2e4*** | 2.37 (1.40, 4.00)** | 2.30 (0.70, 7.53) | 1.96 (0.45, 8.55) |
| ***e3e3*** | 1 (reference) | 1 (reference) | 1 (reference) |
| ***e3e4*** | 3.11 (2.59, 3.72)*** | 2.10 (1.19, 3.70)* | 4.53 (2.52, 8.17)*** |
| ***e4e4*** | 7.00 (5.44, 8.99)*** | 4.54 (1.07, 19.3)* | 7.10 (2.74, 18.4)*** |

Hazard ratios reflecting the risk of amyloid positivity conferred by each *APOE* genotype relative to *e3e3*. For example, *e4e4* carriers were 7.00 times more likely to become A+ compared to *e3e3* carriers in ADNI. A separate Cox proportional hazards model was fitted in each data set. Each model was adjusted for sex. 95% confidence interval for each hazard ratio is indicated in parentheses. * $p<0.05$, ** $p<0.01$, *** $p < 0.001$.

**Supplementary Table 11: Accelerated failure time model outcomes for A+ onset age by *APOE* genotype**

| | Data set | | |
|---|---|---|---|
| ***APOE* genotype** | *ADNI* | *BLSA* | *WRAP* |
| ***e2e3*** | 0.12 (0.06, 0.17)*** | 0.01 (-0.13, 0.15) | 0.08 (-0.09, 0.24) |
| ***e2e4*** | -0.11 (-0.18, -0.04)** | -0.21 (-0.41, -0.01)* | -0.10 (-0.32, 0.12) |
| ***e3e3*** | 0 (reference) | 0 (reference) | 0 (reference) |
| ***e3e4*** | -0.14 (-0.16, -0.12)*** | -0.13 (-0.23, -0.04)** | -0.23 (-0.33, -0.14)*** |
| ***e4e4*** | -0.24 (-0.28, -0.21)*** | -0.28 (-0.57, 0.01) | -0.35 (-0.53, -0.17)*** |

Accelerated failure time model results reflecting the differences in time to A+ onset between each *APOE* genotype and the *e3e3* group. For example, *e4e4* carriers became A+ 24% earlier than *e3e3* carriers in the ADNI. A separate model was fitted in each data set. Each model was adjusted for sex. 95% confidence interval for each hazard ratio is indicated in parentheses. * p<0.05, ** p<0.01, *** p < 0.001.

**Supplemental Table 12: Cox analysis median unimpaired survival and 10-year unimpaired rate for the time from A+ onset to impairment onset**

| | **Median Survival Time Years estimate (95% CI)** | **Ten-year Unimpaired Rate estimate (95% CI)** |
|---|---|---|
| **ADNI Unimpaired Subset (n=595)** | 13.57 (12.98-14.08) | 0.744 (0.703-0.786) |
| **Sex (female>male)** | | |
| Female | 12.89 (11.87-13.66) | 0.699 (0.644-0.755) |
| Male | 14.08 (13.57-14.55) | 0.784 (0.736-0.826) |
| ***APOE*** | | |
| *e4-* | 14.42 (13.97-15.54) | 0.812 (0.766-0.857) |
| *e4+* | 12.73 (11.71-13.42) | 0.691 (0.637-0.741) |
| **EAOA** | | |
| 55 | 14.08 (13.50-14.55) | 0.780 (0.731-0.826) |
| 60 | 14.08 (13.63-14.55) | 0.786 (0.745-0.828) |
| 65 | 13.97 (13.40-14.33) | 0.770 (0.730-0.810) |
| 70 | 13.33 (12.40-13.94) | 0.729 (0.685-0.773) |
| 75 | 11.96 (10.92-13.06) | 0.653 (0.595-0.714) |
| 80 | 10.32 (9.30-11.59) | 0.527 (0.426-0.637) |

Analyses included 595 ADNI participants that were unimpaired prior to A+ onset, excluding *e2e4* carriers (n=15). Covariate adjusted median survival time and ten-year unimpaired rate for time from A+ onset to impairment (i.e., CDR≥1) are shown in the table. No significant interactions were observed between sex and EAOA and between *APOE* and EAOA.
ADNI = Alzheimer's Disease Neuroimaging Initiative, *APOE* = apolipoprotein, CI = confidence interval, EAOA = estimated amyloid onset age

**Supplemental Table 13: Cox analysis median unimpaired survival and 10-year unimpaired rate for the time from A+ onset to impairment onset for *APOE* genotypes**

| | **Median Survival Time Years estimate (95% CI)** | **Ten-year Unimpaired Rate estimate (95% CI)** |
|---|---|---|
| ***APOE*** | | |
| *e3e3* | 14.46 (13.87-15.80) | 0.814 (0.763-0.859) |
| *e3e4* | 13.03 (11.96-13.66) | 0.708 (0.649-0.763) |
| *e4e4* | 11.61 (10.35-13.22) | 0.635 (0.540-0.728) |

Analyses included 595 ADNI participants that were unimpaired prior to A+ onset, excluding *e2e4* carriers (n=15). Covariate adjusted median survival time and ten-year unimpaired rate for time from A+ onset to impairment (i.e., CDR≥1) are shown in the table. No significant interactions were observed between sex and EAOA and between *APOE* and EAOA.
ADNI = Alzheimer's Disease Neuroimaging Initiative, *APOE* = apolipoprotein, CI = confidence interval, EAOA = estimated amyloid onset age

### Supplemental Figures

**Supplemental Figure 1: Inter-method comparison of estimated age A+ a function of the reference DVR/SUVR**

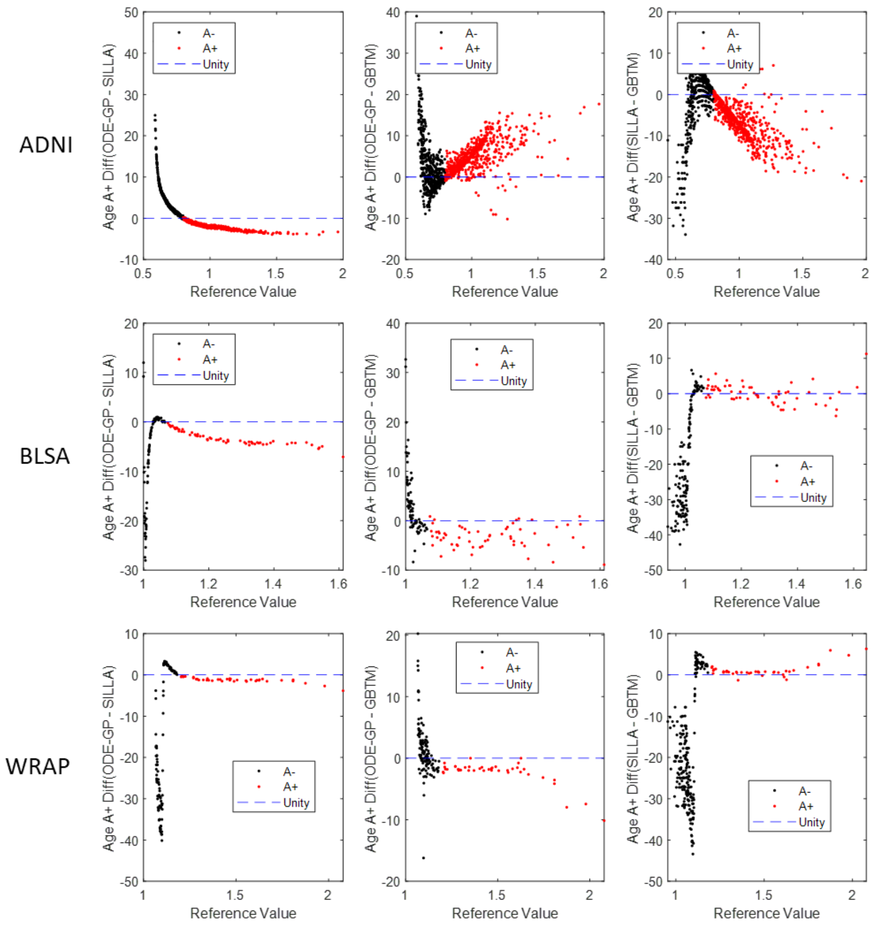


**Supplemental Figure 1:** Comparisons of estimated age A+ between pairs of methods for all participants in each cohort. Age A+ was estimated for all participants in each sample with models trained on the participants in each sample that had two or more amyloid PET scans. Reference value indicates the SUVR (for ADNI) or DVR (BLSA and WRAP) value that was used to estimate each person's age of A+ onset.

**Supplemental Figure 2: EAOA conversion midpoint error in subsets of A- to A+ convertors**

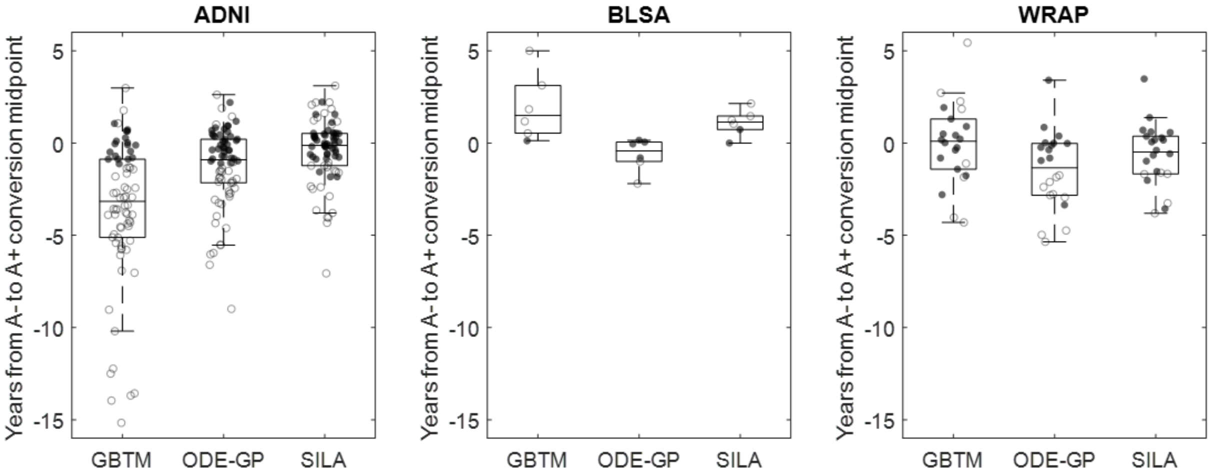


**Supplemental Figure 2:** The time in years from the estimated age of A+ for each method and the midpoint between the last observed A- scan and the first observed A+ scan in the subset of participants from each cohort that were observed to convert from A- to A+ during the course of study (n = 72 for ADNI; n = 6 for BLSA; n = 22 for WRAP). Positive values indicate the model estimated age A+ was greater than the midpoint of the conversion window. Closed circles indicate observations where the model estimated age A+ fell between the last observed A- scan and the first observed A+ scan. Open circles indicate where the model estimated age of A+ was outside of the observed conversion window.

**Supplemental Figure 3: SUVR prediction residuals for ADNI, 10-fold cross-validation**

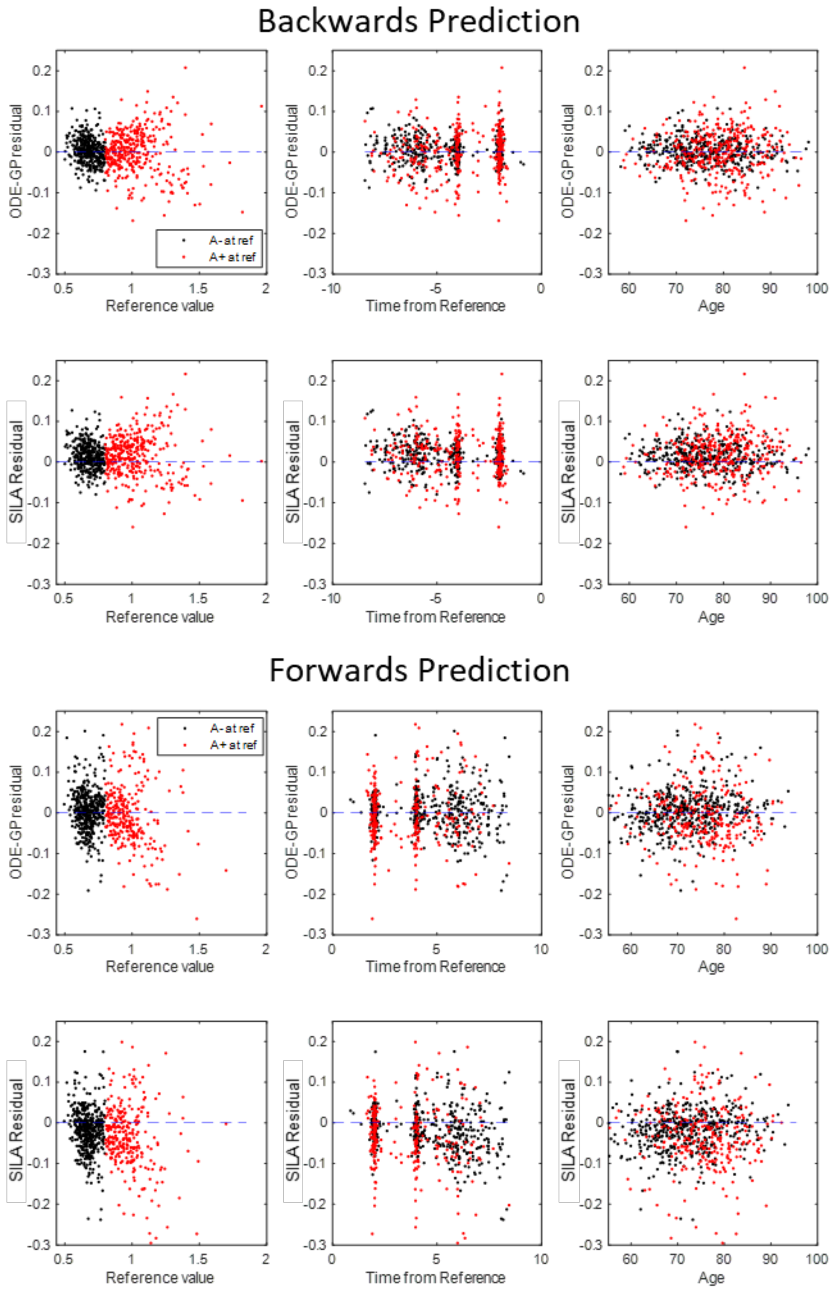


**Supplemental Figure 3:** SUVR prediction residuals for forward and backward prediction for ODE-GP and SILA methods as a function of reference SUVR value, time from the reference scan, and age at the reference scan for ADNI participants with two or more amyloid PET scans. Residuals are calculated using 10-fold cross-validation with either predicting a first scan SUVR from a last reference scan (backward prediction) or predicting a last scan SUVR from a first reference scan (forward prediction).

**Supplemental Figure 4: DVR prediction residuals for BLSA, 10-fold cross-validation**

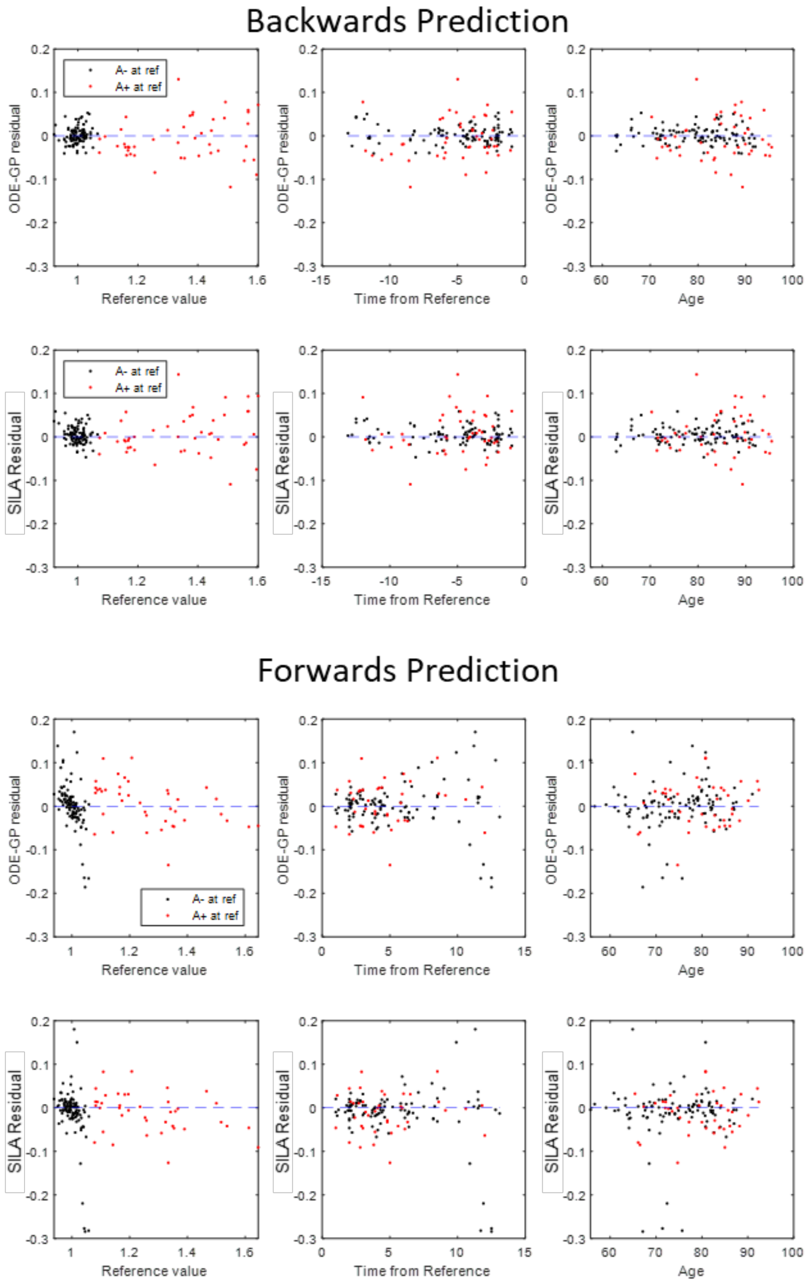


**Supplemental Figure 4:** DVR prediction residuals for forward and backward prediction for ODE-GP and SILA methods as a function of reference DVR value, time from the reference scan, and age at the reference scan for BLSA participants with two or more amyloid PET scans. Residuals are calculated using 10-fold cross-validation with either predicting a first scan DVR from a last reference scan (backward prediction) or predicting a last scan DVR from a first reference scan (forward prediction).

**Supplemental Figure 5: DVR prediction residuals for WRAP, 10-fold cross-validation**

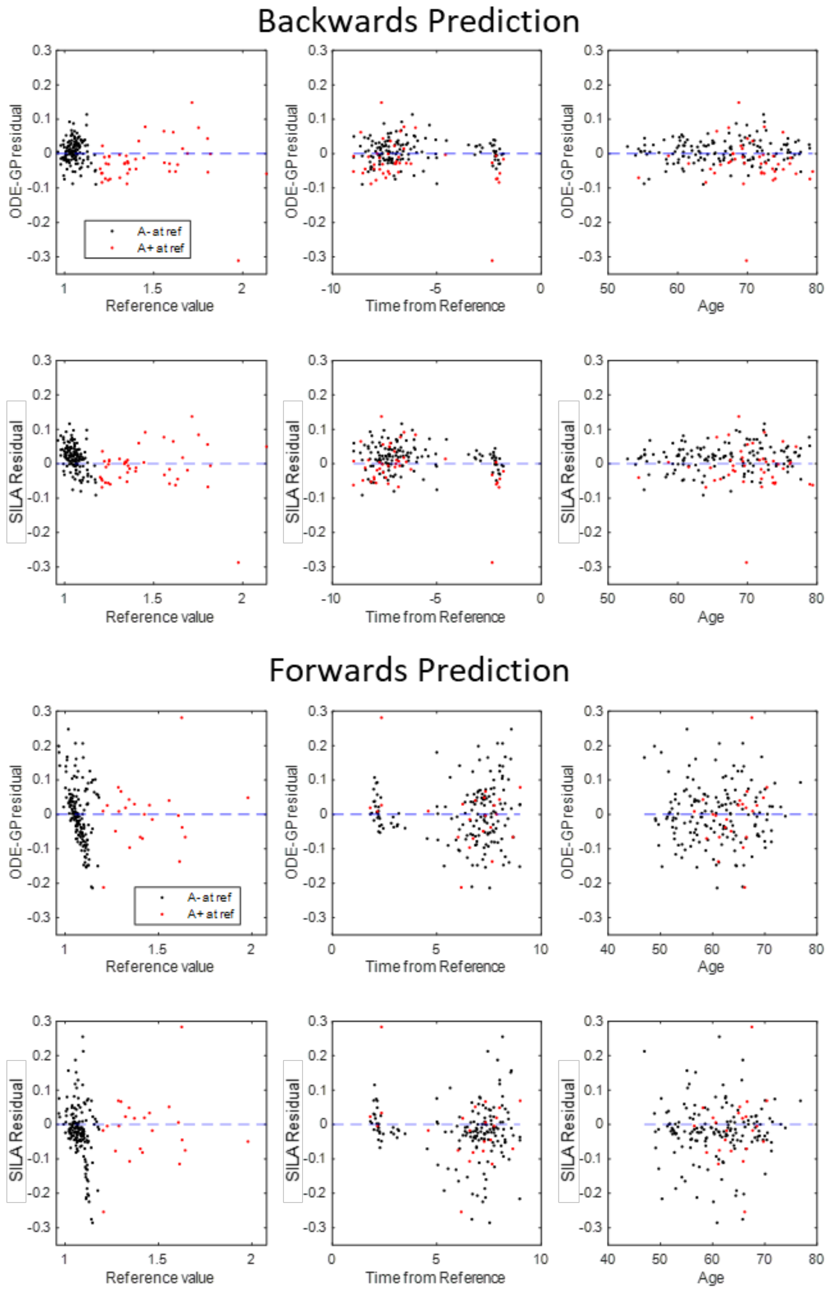


**Supplemental Figure 5:** DVR prediction residuals for forward and backward prediction for ODE-GP and SILA methods as a function of reference DVR value, time from the reference scan, and age at the reference scan for WRAP participants with two or more amyloid PET scans. Residuals are calculated using 10-fold cross-validation with either predicting a first scan DVR from a last reference scan (backward prediction) or predicting a last scan DVR from a first reference scan (forward prediction).

**Supplemental Figure 6: SUVR prediction residuals for ADNI, holdout cross-validation**

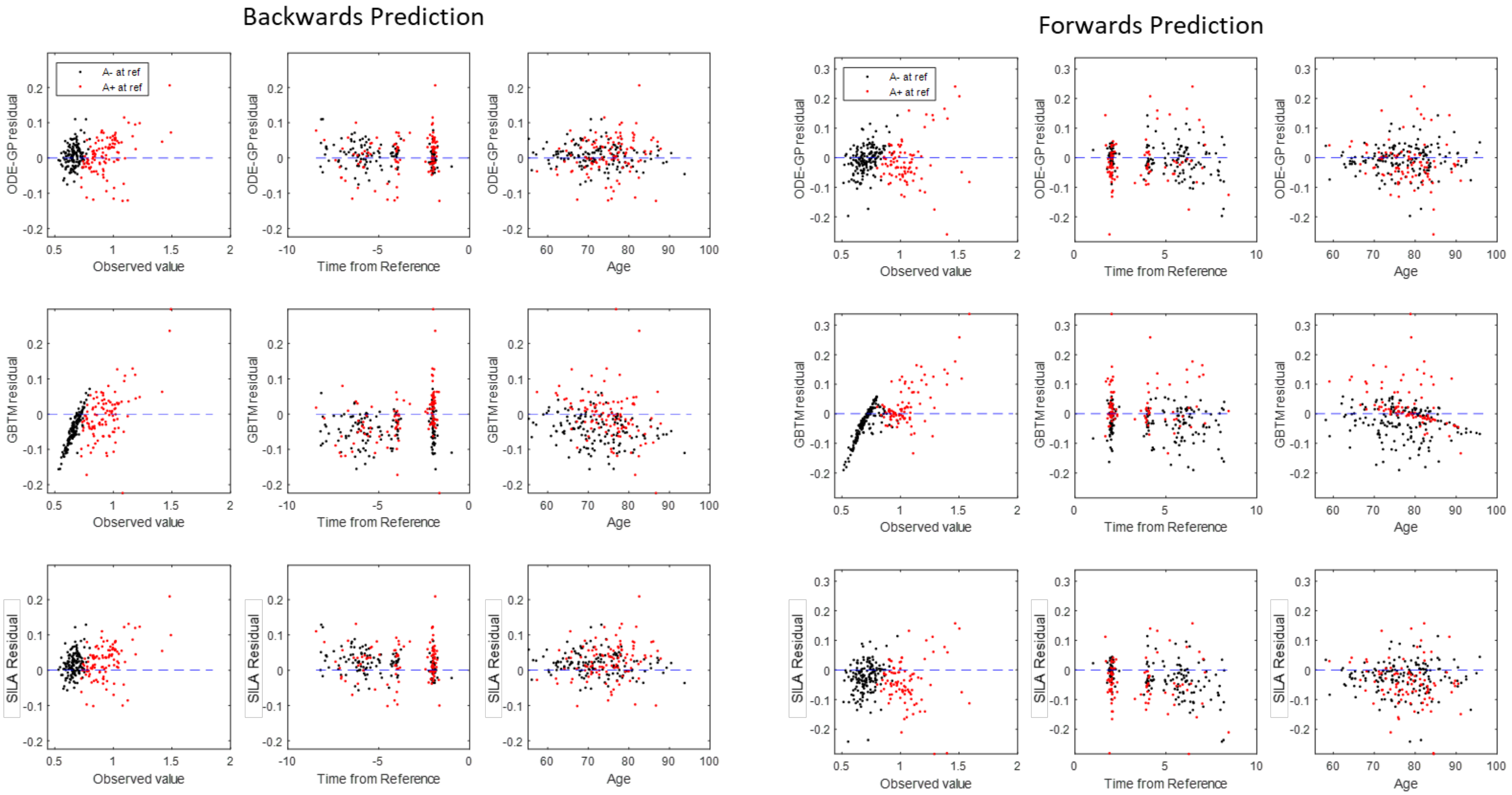

**Supplemental Figure 7: Within-method comparison of estimated age A+ with different model training sets**

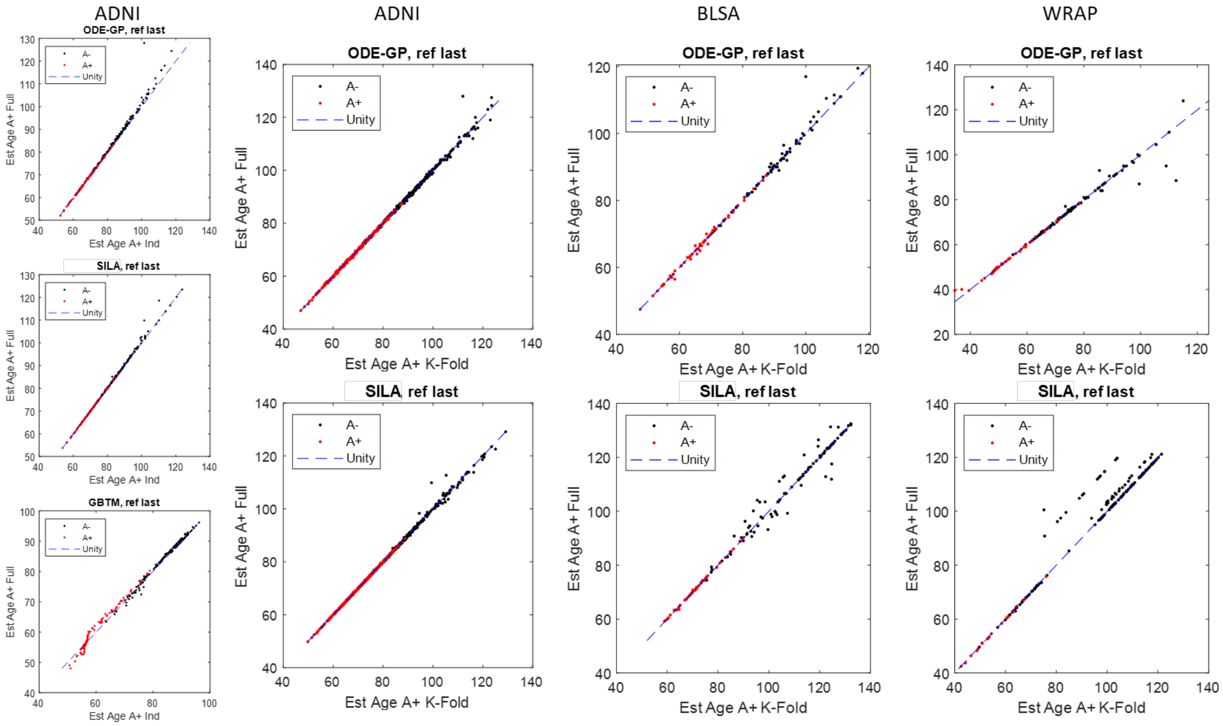

#### Supplemental Figure 8: Amyloid accumulation rates by *APOE4* carriage

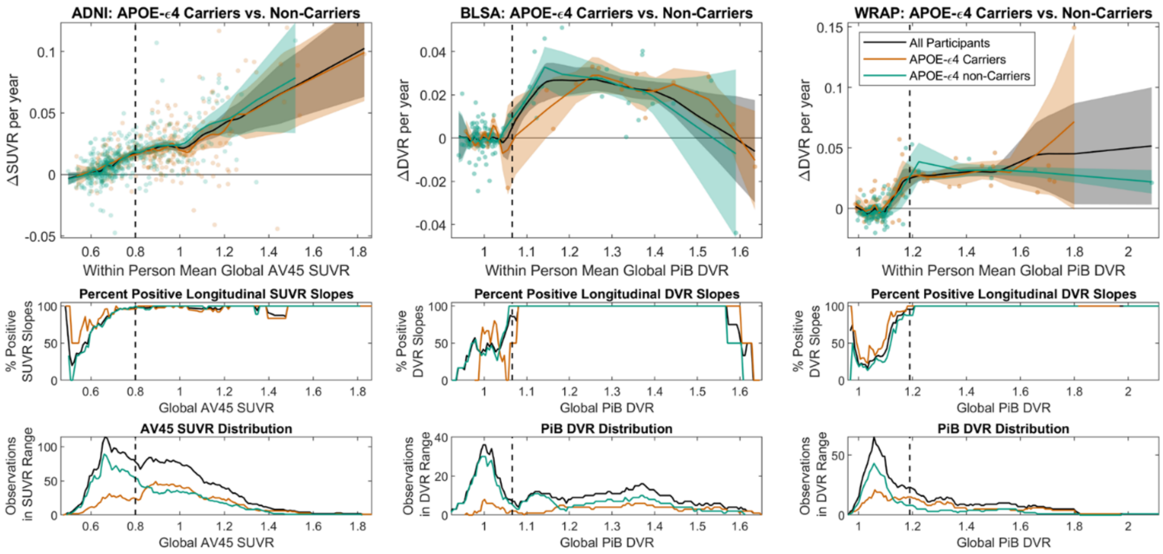

#### Supplemental Figure 9: Amyloid accumulation rates by sex

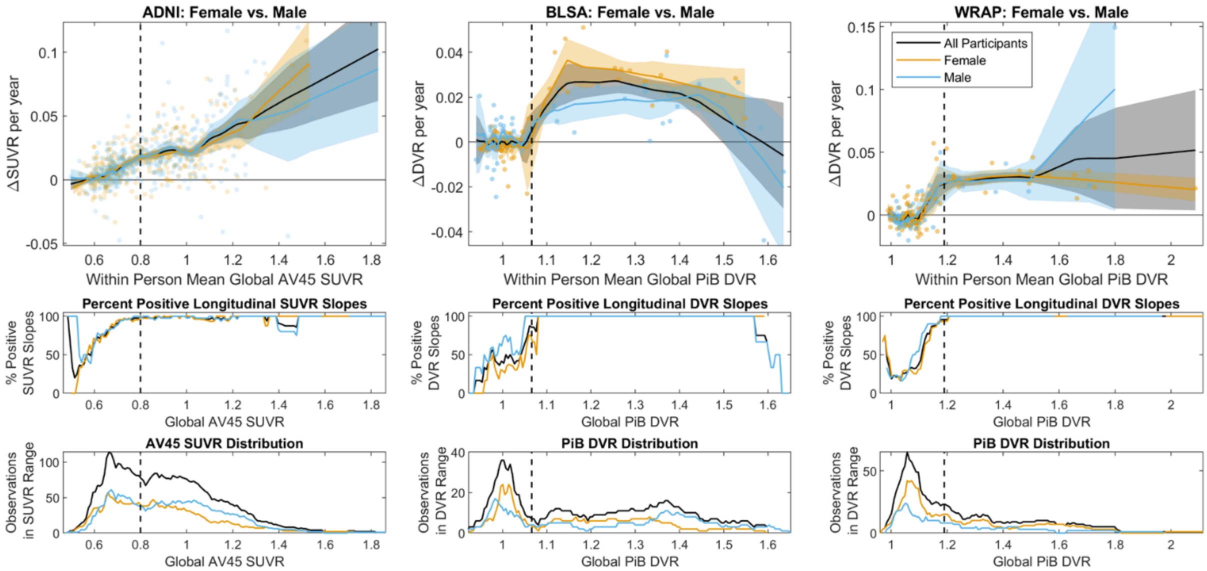

### ODE-GP: A gradient matching technique for fitting univariate ODEs to data

June 16, 2021

#### 1 Statement of the problem

The quantity of interest is univariate and observed over time. We notate this quantity $x(t) \in \mathbb{R}$, with $t \in I$, an interval of times. We assume that $x$ follows a first order ordinary differential equation

$$\frac{\partial x}{\partial t} = \dot{x} = f(x,t), x(t_0) = u, t_0 \in I \tag{1}$$

where $f$ is a regular enough function such that the existence and uniqueness of the solution is guarantied over the interval $I$. We observe $x$ at a collection of time points in $I$, corrupted by noise, and our goal is to estimate $f$ and predict the value of $x$ at arbitrary times. We are interested in the situation where we observe a large number, $n = 100 - 1,000$ trajectories, each with its own initial condition, at a few time points.

$$\begin{cases} \dot{x} & = & f(x,t) \\ x_i(t_0) & = & u_i, i = 1 \dots n \\ z_{ij} & = & x_i(t_{ij}) + \epsilon_{ij}, i = 1 \dots n, j = 1 \dots m_i \end{cases} \tag{2}$$

where $\epsilon$ is a centered random vector assumed to be multivariate normal $MVN(0, \Lambda)$. In the above equation, $z_{ij}$ are observed, $f, u_i$ are unobserved. $\Lambda$ is known or not.

#### 2 Gradient matching

We propose to use a Bayesian method for estimating $f$. Specifically, we assume that $f$ is a smooth Gaussian process, see [1].

$$f \sim GP(\mu, K) \tag{3}$$

Next, we propose to use the numerical scheme known as "gradient matching". It proceeds in two successive steps:

1. Generate pseudo data-points: Estimate $x(t_k)$ and $\dot{x}(t_k, x(t_k))$, as well as error bars at a collection of time points $t_k$
2. Estimate $f$ using Gaussian process regression, [1].

Let us now describe each step of the gradient matching.

#### 2.1 Generate pseudo data-points

This step is data dependent and can be adapted to the situation at hand. In the case of the WRAP, BLSA, or ADNI data, there are a few data points available per subjects, that is $1 \leq m_i \leq 10$. However, the number of subjects is large , with $n > 100$. In this context, we choose to estimate one gradient per subject. We use simple linear regression for this estimate. Thus, for each subject $k$, $k = 1, \ldots, n$,

1. let $t_k = \frac{1}{m_k} \sum_{j=1}^{m_k} t_{kj}$;
2. estimate $x(t_k)$ with $x_k = \frac{1}{m_k} \sum_{j=1}^{m_k} z_{kj}$;
3. $x_k$ is centered at $x(t_k)$ with variance $\frac{\sigma^2}{m_k}$
4. fit the linear model $z_{kj} = a_k t_{kj} + b_k$ using ordinary least squares (OLS);
5. estimate $\dot{x}(t_k)$ with $y_k = a_k$
6. under the standard hypothesis of the ordinary least squares model, $y_k$ is an unbiased estimator of $\dot{x}(t_k)$ with variance $\frac{\sigma^2}{m_k Var(t_{k1}, \ldots, t_{km_k})}$ and independent of $x_k$.

#### 2.2 Estimate $f$ using Gaussian process regression

The model is as follows:

$$\begin{cases} f & \sim & GP(\mu, K) \\ y_i & = & f(x_i + \eta_i, t_i) + \epsilon_i, i = 1 \ldots n \\ \eta & \sim & MVN(0, \Lambda_\eta) \\ \epsilon & \sim & MVN(0, \Lambda_\epsilon) \\ \Lambda_\eta & = & diag(\frac{\sigma^2}{m_i Var(t_{i1}, \ldots, t_{im_i})}, i = 1 \ldots n) \\ \Lambda_\epsilon & = & diag(\frac{\sigma^2}{m_i}, i = 1 \ldots n) \\ (f, \eta, \epsilon) & & \text{are independent} \end{cases} \quad (4)$$

where $MVN$ stands for Multivariate Normal. The goal is to estimate $f$. Unfortunately, this problem is difficult to solve. Indeed, due to the random variables $\eta_i$, $y_i | f$ is not Gaussian and thus classical GP regression cannot be used.

We circumvent this problem as follows. First, we approximate $f$ at $x_i$ by its Taylor extension at order 2:

$$f(x_i + \eta_i, t_i) \approx f(x_i, t_i) + \eta_i f'(x_i, t_i) + \frac{1}{2} \eta_i^2 f''(x_i, t_i) \quad (5)$$

Note that without further approximation $y_i|f$ is still not Gaussian since $\eta_i^2$ is $\chi^2$ distributed. Second, we approximate the right hand side of (5) with its expectation given $f$, that is

$$f(x_i + \eta_i, t_i) \approx f(x_i, t_i) + \frac{1}{2}[\Lambda_\eta]_{ii} f''(x_i, t_i) \tag{6}$$

This method is justified when $f$ is sufficiently regular, see [2], which is a reasonable assumption for the Amyloid accumulation over time. Inference is then performed using the formula for the posterior mean of a MVN distribution

$$E[X_1|X_2 = x_2] = \mu_1 + \Sigma_{12}\Sigma_{22}^{-1}(x_2 - \mu_2) \tag{7}$$

applied to

1. $X_1 = f(u^*)$ of dimension $(m, 1)$;
2. $X_2 = y$ of dimension $(n, 1)$;
3. $\mu_1 = \mu(f(u^*))$ of dimension $(m, 1)$;
4. $\mu_2 = \mu(y)$ of dimension $(n, 1)$;
5. $\Sigma_{11} = K(u^*, u^*)$ of dimension $(m, m)$
6. $\Sigma_{12} = K(u^*, u) + K_{02}(u^*, u)\Lambda_\eta$ of dimension $(m, n)$
7. $\Sigma_{22} = K(u, u) + \frac{1}{2}K_{02}(u, u)\Lambda_\eta + \frac{1}{2}\Lambda_\eta K_{20}(u, u) + \frac{1}{4}\Lambda_\eta K_{22}(u, u)\Lambda_\eta + \Lambda_\epsilon$

with

$$[K_{kl}(u, v)]_{ij} = \frac{\partial^k \partial^l}{\partial^k x_i \partial^l x_j} K(u_i = (x_i, t_i), v_j = (x_j, t_j)) \tag{8}$$

### 3 Choosing $\mu$ and $K$

We choose for $\mu$ a constant function adapted to the data:

$$\mu(x, t) = \frac{1}{n}\sum_{i=1}^{n} y_i \tag{9}$$

and for $K$ the Gaussian kernel

$$K(x_1, x_2) = \exp\left(-(x_1 - x_2)^2/(2\theta^2)\right) \tag{10}$$

The parameter of the kernel is estimated using maximum likelihood.

$$l(\theta) = -\frac{1}{2}\log|\Sigma_{22}(\theta)| - \frac{1}{2}(y - \mu_2)^T \Sigma_{22}^{-1}(\theta)(y - \mu_2) \tag{11}$$

The parameter $\sigma^2$ is estimated using grid search over a held-out data.

#### 4 predictions

Having estimated $f$, we estimate $x(t_1)$, when observing $z(t_0)$ with $t_0 \neq t_1$ using the Euler numerical integration scheme starting at $z(t_0)$.
